# Trans-ancestry genome-wide association meta-analysis of gallstone disease

**DOI:** 10.1101/2025.03.16.25324077

**Authors:** Junghyun Lim, Marijana Vujkovic, Michael G. Levin, Kim Lorenz, Benjamin F. Voight, David Y. Zhang, Max F. Dudek, Matthew C. Pahl, James A. Pippin, Chun Su, Elisabetta Manduchi, Andrew D. Wells, Struan F.A. Grant, Sarah Abramowitz, Scott M. Damrauer, Samiran Mukherjee, Guoyi Yang, David E. Kaplan, Penn Medicine BioBank, Daniel J. Rader

## Abstract

Gallstone disease is a highly prevalent and costly gastrointestinal disease. Yet, genetic variation in susceptibility to gallstone disease and its implication in metabolic regulatory pathways remain to be explored. We report a trans-ancestry genome-wide association meta-analysis of gallstone disease including 88,063 cases and 1,490,087 controls in the UK Biobank, FinnGen, Biobank Japan, and Million Veteran Program. We identified 91 (37 novel) risk loci across the meta-analysis and found replication in statistically compelling signals in the All of Us Research Program. A polygenic risk score constructed from trans-ancestry lead variants was positively associated with liver chemistry and alpha-1-antitrypsin deficiency and negatively associated with total cholesterol and low-density lipoprotein levels among trans-ancestry and European ancestry groups in the Penn Medicine BioBank. Cross-trait colocalization analysis between risk loci and 44 liver, metabolic, renal, and inflammatory traits yielded 350 significant colocalizations as well as 97 significant colocalizations and 65 prioritized genes from expression quantitative trait loci from eight tissues. These findings broaden our understanding of the genetic architecture of gallstone disease.

## INTRODUCTION

Gallstone disease affects 6% of the population globally and generates substantial morbidity, driving one of the largest healthcare costs associated with gastrointestinal diseases.^1,2^ The majority of gallstones (80-90%) are cholesterol stones and the pathogenesis stems from pathways in the metabolism of cholesterol and bile acids.^3,4^ One notable pathway is the cholesterol transport pathway through which cholesterol from peripheral tissues is delivered to the liver where it is secreted into bile.^5^ The formation of gallstones within bile, or lithogenesis, is influenced by shifts in the proportion of bile constituents that lead to supersaturation of biliary cholesterol.^6^ This lithogenesis is governed by the genetic susceptibility and is related to cholesterol homeostasis.^7^

Prior family clustering, twin studies, and genome wide association studies (GWAS) have begun to shine light on the genetic architecture of gallstone disease.^8,9^ Most recently, Fairfield et al. (2022) published a largest gallstone GWAS meta-analysis to date with 43,639 cases and 506,798 controls from the United Kingdom Biobank (UKBB) and FinnGen, identifying 75 risk loci among which 46 were previously unreported.^10^ Previous GWAS have identified a number of associated loci involved in regulatory pathways of cholesterol homeostasis.^4,10–16^ Some of these variants were mapped to genes encoding transporter proteins and genes regulating their expression.^12^ An established gain-of-function missense variant (rs11887534[C]) in *ABCG8* which encodes sterol transporter regulating the intestinal and hepatic efflux of cholesterol is associated with decreased plasma cholesterol and increased biliary cholesterol levels.^17^ Some of the previously discovered genetic loci are linked with other cholestatic disorders (e.g. intrahepatic cholestasis of pregnancy, progressive familial intrahepatic cholestasis, and low phospholipid-associated cholelithiasis) as well as atherosclerotic disease.^18^ As such, the investigation of the genetic basis of gallstone disease has important implications in the pathogenesis of diverse phenotypes that share cholesterol regulatory pathways.

Prior genetic studies have largely focused on single-ancestry populations.^4,10,12–15^ Single ancestry analyses are limited in capturing global genetic diversity: they may fail to uncover genetic signals that are more common in other ancestral groups.^19^ Through the trans-ancestry meta-analysis with the largest sample size reported to date, we leveraged an increased statistical power and genetic variation across ancestry groups to discover novel susceptibility variants and conducted post-GWAS analyses to further characterize their roles in the pathogenesis of gallstone disease.

## METHODS

The discovery cohorts comprised of 88,063 cases and 1,490,087 controls with genotyping data from four cohorts: UKBB,^20^ FinnGen,^21^ Biobank Japan (BBJ),^16^ and Million Veteran Program (MVP).^22^ The cases consisted of individuals of European (EUR) ancestry (72,827 cases; 1,158,486 controls) from UKBB, FinnGen, and MVP, East Asian (EAS) ancestry (9,305 cases; 168,253 controls) from BBJ, African (AFR) ancestry (3,636 cases; 115,711 controls) from MVP, and Admixed American (AMR) ancestry (2,295 cases; 47,637 controls) from MVP. For MVP data, we used summary statistics available from the MVP genome-wide phenome-wide association study (gwPheWAS). The validation set consisted of participants of the All of Us Research Program (AoU)^23^ (5,537 cases; 166,333 controls). Among the cases, 3,294 individuals were of EUR ancestry, 74 of EAS ancestry, and 921 of AFR ancestry. For AoU trans-ancestry group, we included individuals of other ancestry groups (AMR [1,178 cases], South Asian [43 cases], and Middle Eastern [27 cases]) to improve statistical power.

Post-GWAS analyses incorporated participants of Penn Medicine BioBank (PMBB), including 1,628 cases and 37,744 controls. For all cohorts, gallstone disease cases were defined as Phecode 574.1, which corresponds to K80, K80.0, K80.1, K80.2, K80.3, K80.4, and K80.8 for International Classification of Diseases Tenth Revision (ICD-10) codes, and 574.0, 574.00, 574.01, 574.1, 574.10, 574.11, 574.2, 574.20, and 574.21 for ICD Ninth Revision (ICD-9) codes. Controls were all individuals without Phecodes 574-576.99. Cohort-specific descriptions, genotyping platform, imputation, and quality-control are delineated in Supplementary Material and Supplementary Table 1. Of note, we defined EUR, EAS, AFR, and AMR ancestry groups as individuals who are genetically similar to the respective reference populations defined by the 1000 Genomes Project (1kGP) samples.^24^

All aforementioned studies comply with the Declarations of Helsinki and Istanbul and were approved by their respective ethics or institutional review committees: North West - Haydock Research Ethics Committee (UKBB), Ethical Review Board of the Hospital District of Helsinki and Uusimaa (FinnGen), ethics committees of the Institute of Medical Sciences, the University of Tokyo and RIKEN Center for Integrative Medical Sciences (BBJ), the VA Central Institutional Review Board (IRB) (MVP), IRB of the All of Us Research Program (AoU), and IRB of Perelman School of Medicine at the University of Pennsylvania (PMBB).

### Statistical Analysis

We used publicly available summary statistics for UKBB, FinnGen (release 9 data), BBJ, and MVP (gwPheWAS). Genotyped or imputed single nucleotide polymorphisms (SNPs) that met the quality control criteria (imputation quality score [INFO] ≥ 0.3) were included. We excluded SNPs with minor allele frequency (MAF) < 0.01. Genomic coordinates were lifted over to hg38. An inverse variance-weighted, fixed effects model using METAL^25^ was used to perform meta-analysis. In this model, each study’s estimates were weighed by the inverse of their variances (larger sample sizes were alloted more weight) and the effect sizes for all studies were assumed be the same, thereby attributing the difference in the results to random sampling error.^25^ EUR meta-analysis was performed among participants of EUR ancestry from UKBB, FinnGen and MVP, and trans-ancestry meta-analysis was performed among those of EUR ancestry, EAS ancestry from BBJ, AMR ancestry from MVP, and AFR ancestry from MVP. For trans-ancestry and ancestry-specific analyses, we used PLINK 2.0 software^26^ to clump variants using a size of ±500 kb and/or *r*^*2*^ threshold of 0.001 and selected the top associated variant with lowest P-value at each locus as the lead variant. We used ancestry-specific LD reference panels from the 1kGP for respective ancestry-specific analyses and used all individuals from the 1kGP for the LD reference panel for trans-ancestry analysis. The variants were designated genome-wide significant if their P-value was lower than the conventional genome-wide significant P-value threshold of 5×10^−8^. Upon selecting lead variants from each trans-ancestry and ancestry-specific analysis, we again applied the clumping criteria (±500 kb and/or *r*^*2*^ threshold of 0.001) to identify the total number of lead variants across all analyses. Through METAL, we applied Cochrane’s Q test to investigate the heterogeneity of effect sizes between cohorts used in meta-analyses.

The replication study in the AoU evaluated the significance and directional concordance of the trans-ancestry and ancestry-specific lead variants (Supplementary Material). Based on the number of lead SNPs tested, the nominal P-value required for replication was based on Bonferroni-adjustment set to 5.49×10^−4^. We looked up lead SNPs on the Open Targets Genetics^27,28^ and the Ensemble Variant Effect Predictor^29^ to nominate associated genes and annotate the predicted consequence, respectively. Using trans-ancestry summary statistics, we used the causal robust mapping method in meta-analysis (CARMA),^30^ a Bayesian model applied to fine mapping, to ascertain potential causal variants (Supplementary Material).

To quantify the cumulative predictive power of the risk variants, we constructed polygenic risk scores (PRS) using the classic sum of weighted alleles method^31^ allocating weights to each risk allele of lead SNP based on the estimated effects from GWAS (Supplementary Appendix). We then tested risk discrimination for trans-ancestry and ancestry-specific PRS by performing logistic regression for gallstone disease risk and measuring area under the receiver operative curve (AUC) in an independent dataset (PMBB) (Supplementary Material). We next sought to identify additional traits that were associated with the trans-ancestry PRS using a genome-first approach for associations with phenotypes extracted from electronic health record (EHR) among PMBB participants. Extracted phenotypes included phecodes and laboratory measurements (Supplementary Material). A phenome-wide association study (PheWAS) was performed using the “PheWAS” R package. Using logistic regression, we tested associations between each phenotype and trans-ancestry PRS within the trans-ancestry and ancestry-specific groups (Bonferroni-adjusted P-value <2.86×10^−5^). Using this method, we additionally performed PheWAS using the lead, trans-ancestry coding variants (rs11887534[C] at *ABCG8*, rs28929474[T] at *SERPINA1*, and rs1800961[T] at *HNF4A*) in the trans-ancestry group. We then performed linear regression to test associations between each laboratory measurement and the trans-ancestry PRS within the trans-ancestry and ancestry-specific groups (Bonferroni-adjusted P-value <2.78×10^−3^). All PheWAS and laboratory parameter association studies were adjusted for age, genetically inferred sex, and first 10 principal components of genetic ancestry.

To further characterize the genetic architecture of gallstone disease, we estimated SNP-based and tissue-specific heritability using LD score regression (LDSC) (Supplementary Material).^32^ We annotated SNPs to open chromatin regions (OCR), histone mark regions, and promoter interacting regions (PIR) (Supplementary Material). To prioritize potential causal genes at the risk loci, we used colocalization with cross-trait and expression quantitative trait loci (eQTLs) (Supplementary Material). Based on cross-trait colocalization and gene set analysis (Supplementary Material), we associated biological pathways with trans-ancestry lead SNPs and nominated genes, respectively.

## RESULTS

### Genome-wide association meta-analysis

To expand the number of loci associated with gallstone disease, we began by assembling pre-existing and new genome-wide data for disease. Our initial GWAS meta-analysis consisted of 88,063 cases and 1,490,087 controls across four cohorts (UKBB, FinnGen, BBJ, and MVP) incorporating four ancestry groups: EUR (78.0%), EAS (11.3%), AFR (7.6%), and AMR (3.2%). The ancestry group breakdown in each cohort is described in Supplementary Table 1. After meta-analysis of ancestry groups (Methods), we identified 91 loci associated with gallstone disease at genome-wide significance, of which 85 loci were from trans-ancestry group, 4 loci from EUR group, and 2 loci from EAS group (Supplementary Tables 2-5; Figure 1; Supplementary Figures 1-2). Among these loci, 37 loci were previously unreported, of which 33 loci were from trans-ancestry group and 4 loci were from EUR group (Supplementary Tables 2-4). Among novel trans-ancestry lead loci, one locus (rs150754672[T] in *FNDC10*) was found to be genome-wide significant in AFR group (Supplementary Tables 2 and 6; Supplementary Figure 3). We identified no genome-wide significant loci in AMR group and no additional novel loci in EAS group (Supplementary Tables 2 and 5).

**Figure 1.**
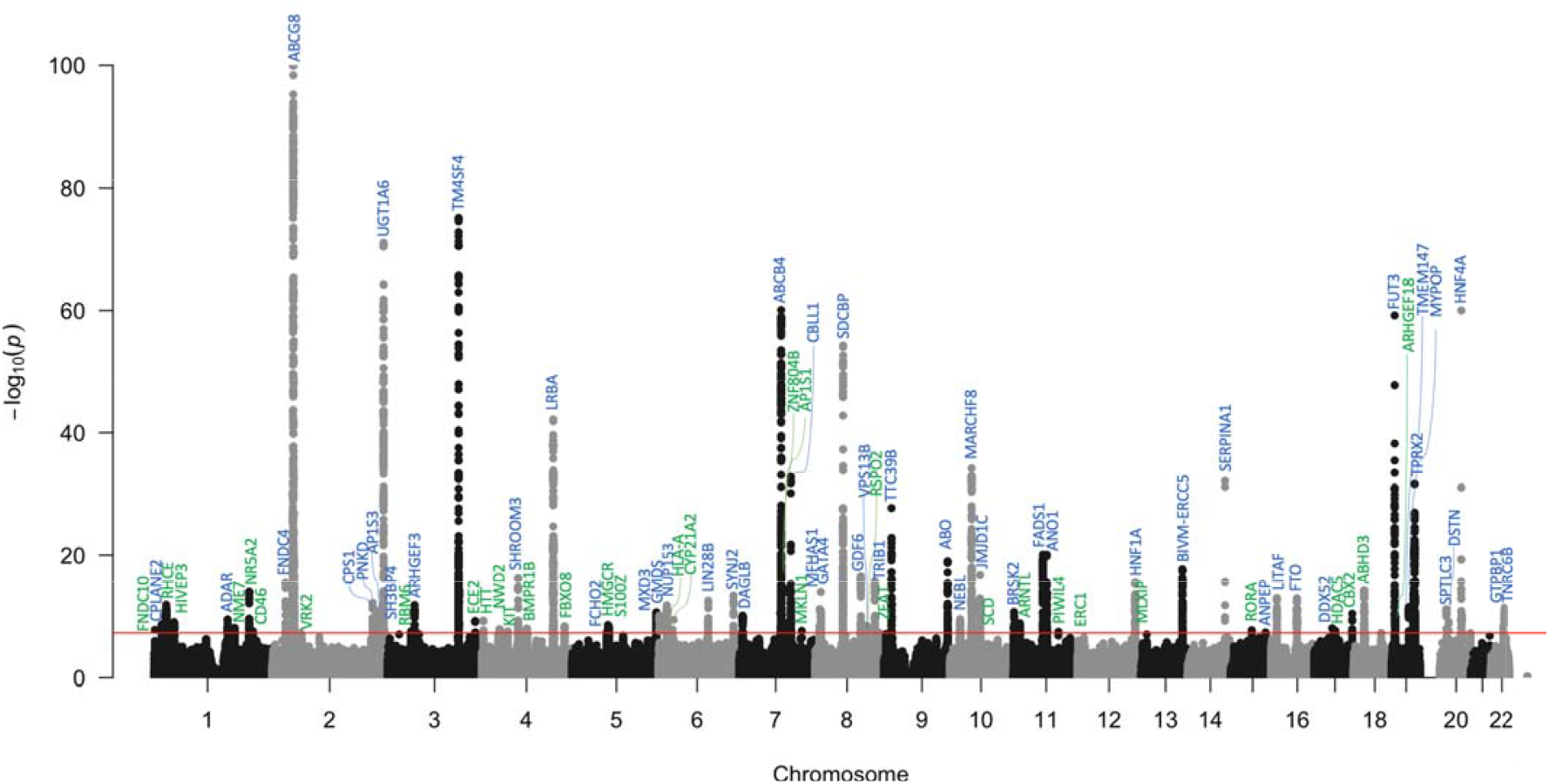
Manhattan plot of genome-wide association in trans-ancestry meta-analysis. (88,063 gallstone cases and 1,490,087 controls). 85 loci met genome-wide significance (p < 5×10^−8^), represented by red line. Novel loci are annotated in green. Known loci are annotated in blue. Y-axis is truncated at p = 10^−100^.

### Replication in All of Us Research Program

We next pursued replication of our sentinel variants in the independent AoU cohort. In a trans-ancestry analysis of AoU (5,537 cases; 166,333 controls), we found 8/85 SNPs exceeded replication significance after Bonferroni correction (P-value <5.49×10^−4^) and 21/85 SNPs exceeded nominal P-value significance (<0.05). Risk alleles at these 21 SNPs were directional consistent between discovery trans-ancestry meta-analysis and AoU. Of these 21 SNPs, one SNP (rs3790848[A] associated with *NR5A2*) was identified as novel in our discovery trans-ancestry cohort. In addition, risk alleles at 67/85 SNPs (79%) were directional consistent between our trans-ancestry meta-analysis and AoU (binomial sign-test P=8.41×10^−8^). The ancestry-specific results of AoU are shown in Supplementary Tables 7-11.

### SNP-based heritability assessment

The SNP-based heritability on the liability scale for gallstone disease was 0.047 (SE 0.0069). Stratified by ancestry groups, the liability-scaled heritability was 0.055 (SE 0.0086) for EUR, 0.075 (SE 0.014) for EAS, and 0.076 (SE 0.048) for AFR (Supplementary Table 12). These estimates were calculated with LDSC (Supplementary Material) using the estimated population prevalence based on Wang et al.^2^

### Fine Mapping

To identify potential causal variants, we performed fine mapping using the CARMA^30^ and constructed credible sets consisting of the smallest number of SNPs with a cumulative posterior probability >95% of containing a true causal variant. Of 85 loci, there was evidence of at least one putative causal variant at 77 loci. Upon selecting SNPs with highest posterior inclusion probability (PIP) within each credible set at each GWAS locus, we identified a total of 129 putative causal SNPs at 77 loci in the trans-ancestry analysis. We looked up the identified SNPs in Open Targets Genetics to learn about the genes. Notably, the locus associated with *ABCG8* was noted to have ten putative causal variants and contains rs11887534[C] which is a known missense variant. In addition, the loci at *TM4SF4* and *ABCB4* had eight putative causal variants at those loci (Supplementary Table 13).

### PRS performance in PMBB

We next tested our gallstone disease PRS in the independent dataset (PMBB) in the trans, EUR, and AFR ancestry groups and assessed the risk discrimination in multivariable-adjusted logistic regression models (Supplementary Table 14; Supplementary Figures 4-5). In the trans-ancestry group, combining trans-ancestry PRS and non-PRS risk factors (described in Supplementary Material) had an improved discrimination accuracy of gallstone disease (AUC 62.7%) compared to non-PRS risk factors (AUC 61.6%). In the EUR group, combining trans-ancestry PRS and non-PRS risk factors (AUC 61.4%) had an improved discrimination accuracy compared to combining EUR PRS and non-PRS risk factors (AUC 61.3%). In the AFR group, combining trans-ancestry PRS and non-PRS risk factors (AUC 63.8%) had an improved discrimination accuracy compared to combining AFR PRS and non-PRS risk factors (AUC 62.8%).

### Cross-phenotypic associations

Using the trans-ancestry PRS, we applied a genome-first approach to investigate the associations with a broad range of phenotypes in PMBB. PheWAS analyses identified seven associations: three in trans-ancestry analysis, three in EUR and one in AFR (Bonferroni significant threshold P <2.862×10^−5^; Supplementary Tables 15-17; Supplementary Figures 6-8). In PMBB trans-ancestry and ancestry-specific groups and as expected, cholelithiasis met the Bonferroni significance. In addition, alpha-1-antitrypsin deficiency met the Bonferroni significance for both trans-ancestry (OR=20.1; P=2.06×10^−13^) and EUR groups (OR=19.9; P=5.29×10^−13^).

We focused our PheWAS analyses in trans-ancestry group on three coding variants identified in trans-ancestry meta-analysis (Supplementary Table 18; Supplementary Figure 9). We tested rs11887534[C] (missense variant in *ABCG8*), rs28929474[T] (missense variant in *SERPINA1*), and rs1800961[T] (missense variant in *HNF4A*). As expected, rs11887534[C] had positive association with cholelithiasis and negative association with hypercholesterolemia. rs28929474[T] had several positive associations, including alpha-1-antritrypsin deficiency, disorders of plasma protein metabolism, emphysema, chronic liver disease and cirrhosis, and liver transplantation. There were no Bonferroni-significant associations for rs1800961[T].

Similarly, the laboratory parameter association study of trans-ancestry PRS was conducted in the trans, EUR, and AFR ancestry groups of PMBB (Bonferroni-significant threshold P <2.778×10^−3^; Supplementary Table 19; Figure 2; Supplementary Figures 10-11). Among Bonferroni-significant associations within the trans-ancestry PMBB group, liver chemistry (aspartate transaminase [AST], alanine transaminase [ALT], total bilirubin [TB], indirect and direct bilirubin) had a positive association and total cholesterol (TC) had a negative association. While LDL did not meet the Bonferroni threshold, it displayed a negative association (Beta=-0.049; P=4.97×10^−3^). Similar findings were observed in the EUR group, where ALT, TB, indirect and direct bilirubin (DB) had a positive association, and LDL had a negative association. TC also showed a negative association (Beta=-0.061; P=3.42×10^−3^) but did not meet the Bonferroni significance. Among the AFR group, TB and indirect bilirubin met the Bonferroni significance and had a positive association. While AST and ALT had a positive association and TC had a negative association, they did not meet the Bonferroni significance.

**Figure 2.**
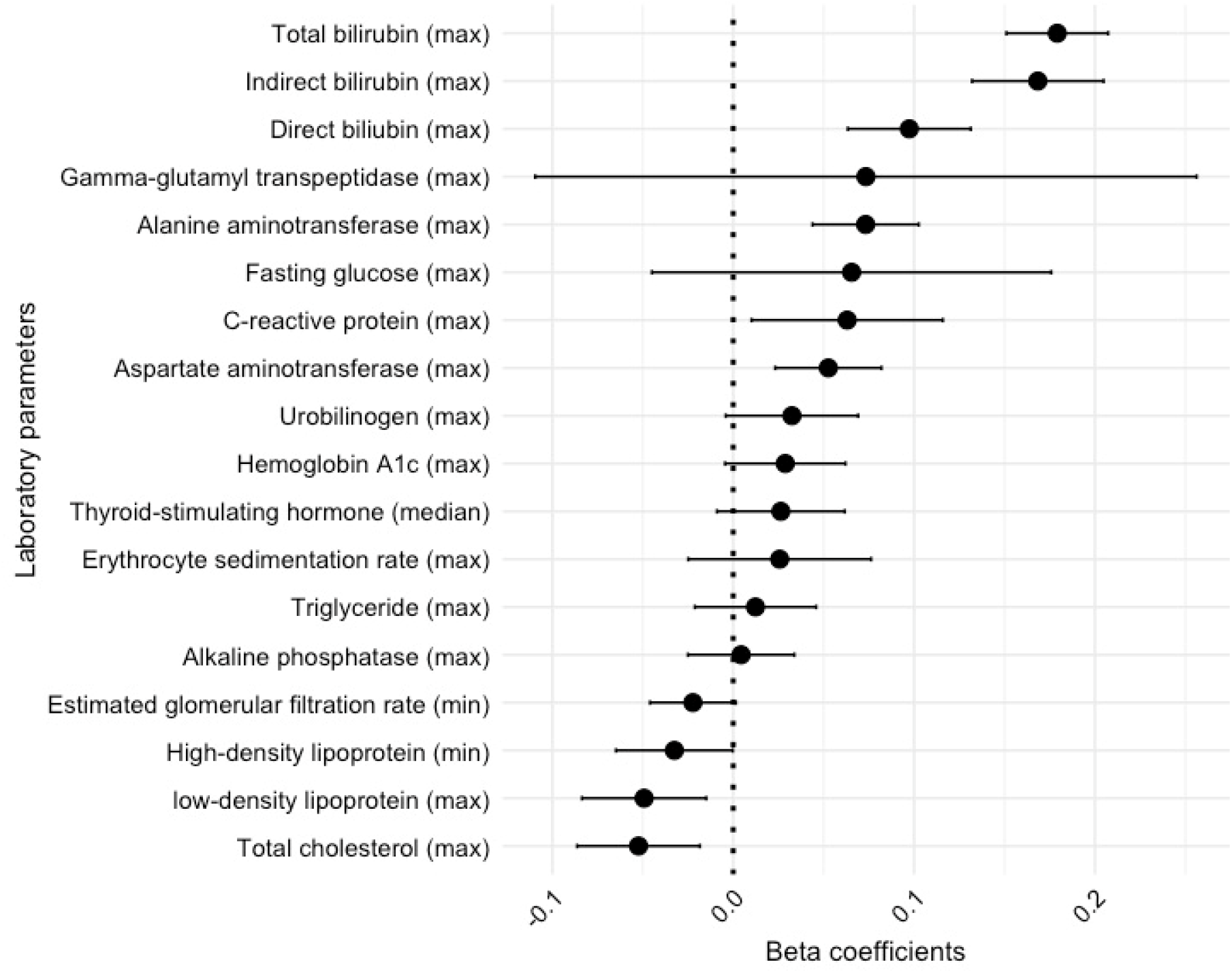
Laboratory parameter association study of the trans-ancestry polygenic risk score in the Penn Medicine BioBank trans-ancestry cohort. Linear regression was used to test the associations between laboratory measurements extracted from the PMBB-linked electronic health record and the trans-ancestry polygenic risk score constructed in PMBB. Bonferroni-significant threshold was set at P <2.778×10^−3^.

### Tissue-specific heritability enrichment

Heritability enrichment analyses were performed for chromatin-based annotations and tissue-specific genes using a false discovery rate threshold of 0.05 for significance (Supplementary Material). When applied to chromatin-based annotations, active enhancer chromatin states in terminal ileum, transverse colon, small and large fetal intestine, and liver demonstrated significant enrichment (Supplementary Table 20). When applied to tissue-specific genes, 53 tissues (including small and large intestine and liver) demonstrated significance (Supplementary Table 21). These findings are consistent with the central role of intestinal and liver tissues in the pathogenesis of gallstones.

### Annotation to chromatin data

We mapped the gallstone susceptibility loci identified via GWAS and fine mapping from trans-ancestry meta-analysis to open chromatin regions (OCR), histone marks, and promoter interacting regions (PIR) in liver, colonoids, and pancreatic tissues. Specifically, OCR, histone marks, and PIR from liver cancer cell line (HepG2) cells, OCR and PIR from hepatocyte-like cells derived from induced pluripotent stem cells, and OCR from human liver cells were used for this purpose. The loci were mapped to OCR and PIR from colonoids and alpha and beta cell types from pancreatic tissues and OCR were mapped to delta, ductal, endothelial, epsilon, mesothelial, and pancreatic polypeptide cell types from pancreatic tissues (Supplementary Table 22; Supplementary Figures 12-14). This annotation provides insights into the potential role of susceptibility loci in the hepatic, colonic, and pancreatic cell types. Further research is needed to elucidate the biological implications of these annotations in the pathogenesis.

### Cross-trait and eQTL colocalization

To further characterize the pleiotropy of the gallstone loci, we analyzed GWAS of 44 related liver, metabolic, renal, and inflammatory traits with our gallstone disease trans-ancestry lead loci and identified 350 significant colocalizations (Supplementary Table 23). Among colocalizations, 42 loci were associated with liver traits, 51 with metabolic traits, 13 with renal traits, and 11 with inflammatory traits. In addition, we tested colocalization for lipoprotein, TB, and DB with all gallstone loci with P-value greater than 5×10^−8^ and less than 5×10^−5^ which identified another 16 significant colocalizations (Supplementary Table 24).

Using the trans-ancestry lead loci, we performed colocalization with eQTLs in eight tissues using GTex summary statistics (Supplementary Material), which yielded 97 significant colocalizations (Supplementary Table 25). In the analysis, we prioritized 65 genes, including 23 expressed in liver, 25 in colon, 14 in spleen, eight in small intestine, and eight in pancreas. One notable gene is *SPTLC3* expressed in liver with a significant colocalization for LDL trait with negative effect direction (Supplementary Table 26).

*SPTLC3* is known to encode a subunit of serine-palmitoyltransferase involved in sphingolipid biosynthesis, potentially implicating a role of sphingolipids in gallstone development.^33^ When we performed colocalization with eQTL in eight tissues for LDL, TB and DB traits for gallstone loci with P-values less than 5×10^−5^, we did not identify additional significant colocalizations (Supplementary Tables 26-28). We note that for loci with numerous credible sets (such as the locus at rs11887534), colocalization analysis is not valid. Given the colocalization analysis is a “rule-in” test, the absence of significant colocalizations with rs11887534 (for instance, with LDL) does not exclude the potential colocalizations.

### Pathway Analysis

In cross-trait colocalization analysis, 25 loci colocalized with only lipid traits, 5 loci with only bilirubin traits, and 10 loci with both lipid and bilirubin traits (Supplemental Table 29), suggesting biological pathways related to lipid metabolism and cholestasis. 45 loci did not colocalize with either traits. In gene set analysis, 54 nominated genes were associated with biological pathways defined by KEGG, Reactome, Wikipathways, and Gene Oncology (Supplemental Table 30). Specifically, 18 genes were associated with lipid metabolism pathway, 3 genes with cholestasis pathway, 3 genes with pathway related to maturity-onset diabetes of the young, and 4 genes with cardiac progenitor differentiation pathway. Most of these genes were also associated with broader homeostatic metabolic process involving a range of cellular functions.

## DISCUSSION

We report 91 (37 novel) genome-wide significant gallstone-susceptibility loci identified across trans, EUR, EAS, and AFR-ancestry meta-analyses. We identified four novel genome-wide significant variants (non-coding variants in *NIPAL1, TARDBPP5, GLIS3*, and *NPC1*) unique to EUR-specific analysis. Among novel trans-ancesty lead loci, one non-coding variant in *FNDC10* was genome-wide significant in AFR-specific analysis. Fine mapping efforts in trans-ancestry meta-analysis yielded 129 putative causal variants at 77 loci, including three missense variants (rs4148323[A] in *UGT1A1*; rs8187801[T] in *ABCB4*; rs56398830[A] in *SLC10A2*). rs8187801[T] and rs56398830[A] were previously reported to be associated with gallstone disease^12,34^ and rs4148323[A] was associated with bilirubin levels.^35^ Overall, these results provide new insight into the genetic architecture of gallstone disease and shared pathways with other metabolic diseases.

Among novel loci, we note the locus located at *NR5A2* associated with an increased risk of gallstone disease in trans-ancestry (rs3790848[A]; effect allele frequency (EAF): 0.36; predicted consequence: intronic) and EUR ancestry (rs28698778[G]; EAF: 0.32; predicted consequence: intergenic). These two variants are located 15,424 base pairs apart, thus rendered LD dependent and reported as one lead locus across all analysis. *NR5A2* encodes a nuclear receptor (NR5A2, or liver receptor homolog 1 [LRH-1]) that is expressed highly in the liver and regulates gene expression in the pathways of cholesterol and bile acid metabolism.^36^ LRH-1 regulates cholesterol transport by binding to an intergenic region in *ABCG5/ABCG8* and stimulating their promoter activity.^36,37^ LRH-1 also influences bile acid homeostasis by mediating *CYP7A1* (encodes rate limiting enzyme catalyzing cholesterol conversion to primary bile acids) and *CYP8B1* (encodes enzyme partaking in cholic acid synthesis).^38,39^ Loss-of-function of LRH-1 is known to decrease *CYP8B1* expression and cholic acid levels, affecting the hydrophobicity of bile.^36^ In addition, LRH-1 activates the promoter of bile salt export pump (BSEP) which mediates the secretion of bile salts from hepatocytes into the biliary system.^36,40^ This is consistent with the pathway analysis, in which we associated *NR5A2* with lipid metabolism pathway. In Capture-C data, we mapped *NR5A2* locus to a promoter interacting region in hepatocyte-like cells and colonoids and identified its interacting gene as *NR5A2*. Colocalization analysis to examine the pleiotropy of this locus did not reveal associated traits. The biological mechanism of the *NR5A2* locus in altering the complex function of the LRH-1 leading to gallstone disease warrants further research.

The laboratory parameter association study of trans-ancestry PRS showed expected findings of a positive association with liver chemistry and a negative association with TC and LDL levels (Supplementary Table 20; Figure 2). These findings highlight the interplay between genetics of gallstone disease and cholesterol homeostasis. For instance, *ABCG8* D19H, an established gain-of-function variant, is associated with decreased LDL, decreased risk of myocardial infarction, and increased risk of gallstone disease.^7^ Among novel loci identified in our trans-ancestry analysis, rs339969[A] in *RORA* (predicted consequence: intronic) showed a positive association in GWAS and a negative association with LDL in colocalization. *RORA* encodes retinoic acid receptor-related orphan receptor-α (RORα) which has been shown to suppress transcriptional activity of PPARγ.^41^ Activation of PPARγ modifies cholesterol and bile acid transporter gene expression, which leads to the net effect of increased biliary bile acid, increased fecal cholesterol and decreased fecal bile acids.^42^ By inhibiting PPARγ-mediated transcriptional cascade, RORα may promote pathways leading to gallstone disease, raising the question whether RORα can be utilized as a preventive target. As such, the investigation of genetic architecture of gallstone disease has important implications in the elucidation of cholesterol regulatory pathways and may advance pharmacogenomics in the disease management.

A genome-first approach to characterize pleiotropy further revealed strong positive association with alpha-1-antitrypsin deficiency (AATD) for trans-ancestry PRS among trans-ancestry and EUR-ancestry groups (Supplementary Figures 6-7). This appears to be driven by the large effect of rs28929474[T] in *SERPINA1* which is an established missense variant^12^ identified as a genome-wide significant locus in our trans and EUR ancestry meta-analysis, consistent with findings by Ferkingstad et al.^12^ This missense variant, known as the Protease Inhibitor (PI) Z allele, leads to altered folding of alpha-1-antitrypsin protein that builds up in the endoplasmic reticulum and causes chronic liver injury, fibrosis, and cirrhosis.^43^ Cross-trait colocalization analysis for this variant showed a positive effect direction with liver traits (e.g. cirrhosis, liver chemistry measures [AST, ALT, ALP, gamma glutamyl transpeptidase, and DB]) and a negative effect direction with coronary artery disease, type 2 diabetes, and C-reactive protein.

The pathogenesis of gallstone disease in AATD is unclear. The changes in physiology (e.g. hypersplenism and increased hemolysis; low bile salt to unconjugated bilirubin molar ratio) in cirrhosis may predispose gallstone development;^44^ however, the underlying mechanism specific to the etiology of AATD remains to be explored.

We note following limitations of the study. Without universal screening for gallstones, we recognize that it is not possible to capture all gallstone cases especially those who are asymptomatic. Thus, asymptomatic individuals with undiscovered gallstones may have been misclassified as controls. In addition, when asymptomatic gallstones are incidentally discovered, discrepancies may exist at the clinician level with translation into gallstone ICD codes. We expect that misclassification of phenotypes would lead to a bias in favor of the null hypothesis. We also acknowledge the potential bias in discovery of asymptomatic gallstones, as those with more complex medical comorbidities may have undergone more imaging studies and be more likely to exhibit incidentally discovered gallstones. Another notable limitation is the lack of data on gallstone types (cholesterol, pigment, or mixed). While the majority of gallstones are cholesterol stones, inability to distinguish the composition leads to limitations in interpreting the biological basis of discovered loci. To address this limitation, we performed a series of post-GWAS analyses including colocalization to gain an insight into their role in gallstone pathogenesis.

While we report the largest sample size published to date in our discovery GWAS meta-analysis, we acknowledge the small sample size of (1) non-European ancestry compared to European ancestry in the discovery cohort, (2) AoU replication cohort, and (3) PMBB for PheWAS. Insufficient statistical power may explain the absence of genome-wide significant loci in AMR group or novel loci in EAS group in our discovery analyses. As most of the lead SNPs were identified in the trans-ancestry and EUR groups, it is plausible that the identification was driven more by the increased sample size than the addition of non-European individuals. While we find the significance of directional concordance with lead SNP risk alleles in the AoU cohort compelling, its small sample size may explain the low rate of replication at the Bonferroni significance threshold. In this context, we note a novel lead locus (rs150754672[T] at *FNDC10*) in trans-ancestry and AFR group which did not meet nominal P-value threshold in AoU and the direction of effect was discordant in trans-ancestry group and concordant in AFR group; therefore, validity of this locus may be questioned. In post-GWAS analyses, we acknowledge that the PheWAS would benefit from incorporating a cohort with a larger sample size. Moving forward, future research with a greater non-European ancestry sample size is warranted to better capture the variability in the human genome, such as those that are shared between ancestry groups, unique to ancestry groups, and the differences in linkage disequilibrium (LD) structures between groups.^45^

In conclusion, we performed a large-scale trans-ancestry genome-wide meta-analysis of gallstone disease using UKBB, FinnGen, BBJ, and MVP, and discovered 91 (37 novel) gallstone-susceptibility loci. We replicated our findings in the AoU and conducted a series of downstream analyses aimed to enhance our understanding of the genetic landscape of gallstone disease. The investigation of genetic basis of bile lithogenicity has the potential to advance our understanding of pathogenesis and underlying regulatory mechanisms of gallstone disease and other diseases that share the biological pathways.

## Data Availability

Summary statistics will be available for download at: https://api.kpndataregistry.org/api/d/CsBP8L
GEO
New HepG2 ATAC/CaptureC: GSE262484
HepG2 ChIP: GSE274031 (reviewer token: ytmfiueqhpqtngz)
HLC ATAC: GSE274004 (reviewer token: ynwdqawmnrcbnyv)
HLC Capture-C: GSE189026
Pancreas Hi-C: GSE188311
HPAP
HPAP040 RRID: SAMN19776471
HPAP045 RRID: SAMN19776476
HPAP054 RRID: SAMN19776484
HPAP066 RRID: SAMN19842595
HPAP072 RRID: SAMN19842601

## RESOURCE AVAILABILITY

### Data and code availability

Summary statistics will be available for download at:

https://api.kpndataregistry.org/api/d/CsBP8L

<u>GEO</u>

New HepG2 ATAC/CaptureC: GSE262484

HepG2 ChIP: GSE274031 (reviewer token: ytmfiueqhpqtngz)

HLC ATAC: GSE274004 (reviewer token: ynwdqawmnrcbnyv)

HLC Capture-C: GSE189026

Pancreas Hi-C: GSE188311

<u>HPAP</u>

HPAP040 RRID: SAMN19776471

HPAP045 RRID: SAMN19776476

HPAP054 RRID: SAMN19776484

HPAP066 RRID: SAMN19842595

HPAP072 RRID: SAMN19842601

## ACKNOWLEDGMENTS

We acknowledge the Penn Medicine BioBank (PMBB) for providing data and thank the patient-participants of Penn Medicine who consented to participate in this research program. We would also like to thank the Penn Medicine BioBank team and Regeneron Genetics Center for providing genetic variant data for analysis. The PMBB is approved under IRB protocol# 813913 and supported by Perelman School of Medicine at University of Pennsylvania, a gift from the Smilow family, and the National Center for Advancing Translational Sciences of the National Institutes of Health under CTSA award number UL1TR001878. We gratefully acknowledge All of Us participants for their contributions, without whom this research would not have been possible. We also thank the National Institutes of Health’s All of Us Research Program for making available the participant data examined in this study. We would like to thank Mitch Connery for post-processing the HepG2 ChIP-seq data. JL was supported by the NHGRI under award 5T32HG009495. MGL was supported by the Doris Duke Foundation (Award 2023-0224) and US Department of Veterans Affairs Biomedical Research and Development Award IK2-BX006551. DYZ was supported by NHLBI under the award number F30HL172382. BFV acknowledges support from the NIH/NIDDK (UM1DK126194).

## AUTHOR CONTRIBUTIONS

JL, MV, MGL, and DJR designed the study. JL, MV, MGL, KL, DYZ, MFD, SA, GY, and SM analyzed data. MFD annotated the SNPs with chromatin data and performed the heritability enrichment analysis. MCP helped to curate the chromatin annotation data and methods. JAP generated the HepG2 ATAC, Capture-C, and ChIP-seq data. CS processed the colonoid ATAC-seq and HepG2 datasets. EM processed the colonoid Capture-C and HLC datasets. JL, MV, MGL, KL, BFV, DYZ, MFD, SD, GY, and DJR interpreted data. JL, MV, KL, DYZ, MFD, MCP, JAP, GY, and SA wrote the manuscript. JL, MV, MGL, KML, BFV, DYZ, MFD, MCP, CS, EM, ADW, SFAG, DEK, GY, and DJR reviewed and edited the manuscript. DJR supervised the study.

