## Supplementary Methods and Figures for "Trans-ancestry genome-wide association meta-analysis of gallstone disease"

### Supplemental Material

#### Contents:

1. Cohort descriptions
2. Replication study
3. SNP-based liability-scaled heritability
4. Polygenic risk score construction and predictive performance
5. Ancestry-specific principal components in Penn Medicine BioBank
6. Phenotypes extracted for phenome-wide and laboratory parameter association study
7. Fine Mapping
8. Tissue-specific heritability enrichment analysis
9. Annotation to chromatin data
10. Cross-trait colocalization
11. Expression quantitative trait locus (eQTL) colocalization
12. Gene set analysis
13. Supplemental figures
14. Penn Medicine BioBank Banner Author List and Contribution Statements
15. References

#### 1. Cohort descriptions

UK Biobank

The UK Biobank (UKBB) is a prospective cohort consisting of approximately 500,000 United Kingdom individuals who were 40-69 years of age at the time of recruitment (2006-2010).[1] Detailed cohort characteristics[2] and description of genotyping platforms and imputation are described elsewhere.[3] We accessed the publicly available summary statistics containing genotyping and phenotyping data (https://docs.google.com/spreadsheets/d/1AeeADtT0U1AukliiNyiVzVRdLYPkTbruQSk38DeutU8/edit?gid=1450719288#gid=1450719288), and included 17,278 cases and 399,542 controls of European ancestry for GWAS meta-analysis. Gallstone disease cases were defined as Phecode 574.1 for all cohorts described in this section and below.

FinnGen

The FinnGen is a public-private partnership established in 2017 which is comprised of approximately 500,000 participants of Finnish biobanks and incorporates genomic data with the clinical data from Finnish national health registries.[4] Mean age at the time of DNA collection was 51.8 years old. Further information regarding genotyping, imputation, and quality control are described previously.[5] We accessed summary statistics of FinnGen release 9 (https://console.cloud.google.com/storage/browser/finngen-public-data-r9/summary_stats/) and included 37,041 cases and 330,903 controls of European ancestry.

Biobank Japan

The Biobank Japan (BBJ) is a large prospective cohort that is nation-wide and hospital-based, consisting of approximately 200,000 individuals from 66 hospitals, recruited from June 2003 and March 2008. Mean age at the time of recruitment was 63 years. DNA samples were collected at baseline and annual serum samples were collected until 2013. Detailed information including the baseline demographic data, collection of deep phenotype data, and genotyping/quality control method are described previously.[6,7] We accessed summary statistics (https://pheweb.jp/pheno/Cholelithiasis) and included 9305 cases and 168,253 controls of East Asian ancestry.

Million Veteran Program

The Million Veteran Program (MVP) is a nation-wide biobank that was established in the Department of Veterans Affairs (VA) health care system under the United States Veterans Health Office of Research and Development. The enrollment of participants began in 2011 and blood samples and deep phenotype data were collected. Further information regarding the baseline demographic data, study structure and design, and genotyping/quality control method are described previously.[8,9] For the analyses, we used summary statistics available from the MVP genome-wide phenome-wide association study (gwPheWAS). We included 18,508 cases and 428,041 controls of European ancestry, 2,295 cases and 47,637 controls of Admixed American ancestry, and 3,636 cases and 115,711 controls of African ancestry.

All of Us Research Program

The All of Us Research Program is a longitudinal cohort based in the United States and collects biospecimens and clinical data from participant surveys and electronic health record from the health care organizations funded by All of Us across the nation. Details regarding the participant recruitment, biospecimen collection, genome sequencing, and quality-control methods are described in prior literature.[10,11] We used the genomic and phenotyping data to replicate our findings from trans-ancestry and ancestry-specific GWA meta-analyses.

Penn Medicine BioBank

Penn Medicine BioBank (PMBB) is a biobank comprising of biospecimen from patients of the University Pennsylvania Health System and is linked to the clinical data from the hospital system's electronic health record (https://pmbb.med.upenn.edu/biobank.php). Approximately 45,000 participants have undertaken genotyping and whole exome sequencing, through the support of the Regeneron Genetics Center. Further information on genotyping platforms and imputation method are described elsewhere.[12] We used the genotyping and phenotyping data from PMBB for post-GWAS analyses.

**2. Replication study**

This study used data from the All of Us Research Program's Controlled Tier Dataset v7, available to authorized users on the Researcher Workbench. From the All of

Us Research Program, we interrogated 245,460 individuals with short-read whole-genome sequencing including 125,860 individuals of European ancestry, 50,080 individuals of African ancestry, and 7,440 individuals of East Asian ancestry. Genetically inferred population assignments provided by the All of Us Research Program were used to subset the cohort for performing downstream analyses. Ancestry-specific subcohorts were categorized as those of European, African, and East Asian ancestry and trans-ancestry subcohort included individuals of all ancestry groups. ICD-9CM and ICD-10CM diagnosis codes available up to July 2022 were extracted from the electronic health record and individually mapped to phecodes.[13] Gallstone cases were defined as individuals with at least two or more dates of Phecode 574.1 and controls were defined as those without the corresponding phecode.

Genome-wide significant loci identified in trans-ancestry and ancestry-specific GWA meta-analyses (See Supplementary Appendix 4) were assessed among the cases and controls of participants of the All of Us Research Program. All associations with cases were performed using generalized linear mixed models as implemented by SAIGE[14], adjusting for sex, age, $age^2$, and the first 10 PCs. Sex and PCs were provided by the All of Us Research Program, and sex was defined as “biological sex assigned birth.” SAIGE estimates both random and fixed effects: random effects include any variation in the phenotype that may be not captured by fixed effects. Based on the number of lead loci tested, Bonferroni corrected P-value ($5.49 \times 10^{-4}$) was used to define significance. Concordance of the effect directionality was compared using the exact binomial test (Supplemental Table 10).

**3. SNP-based liability-scaled heritability**

The “MungeSumstats” R package (https://github.com/neurogenomics/MungeSumstats) and the “ldscr” R package (https://mglev1n.github.io/ldscr/) were applied to perform the LD score regression (LDSC) using ancestry-specific LD reference files from Pan-UK Biobank.[16] Sample and population prevalence[17] were used to derive the heritability coefficients on the liability scale (Supplementary Table 11).[18]

**4. Polygenic risk score construction and predictive performance**

The lead susceptibility variants identified in trans-ancestry and ancestry-specific GWAS were used to construct the trans-ancestry and ancestry-specific polygenic risk score (PRS) by allocating weights to each risk allele based on the risk allele effects estimated in the corresponding GWAS. The PLINK 2.0 software was used to calculate the weighted PRS.

The PRS performance was assessed using risk prediction model for gallstone disease (case defined as individuals with at least two or more dates of Phecode 574.1 and controls defined as those without the corresponding phecode [574-576.99]) in trans-ancestry and ancestry-specific groups in PMBB. Non-PRS risk factors used to adjust models consisted of age, genetically inferred sex, body mass index [median], and first 10 principal components of genetic ancestry. In trans-ancestry group, we tested two logistic regression models: (1) non-PRS risk factors and (2) trans-ancestry PRS + non-PRS risk factors. In ancestry-specific groups, we tested three logistic regression models: (1) non-PRS risk factors, (2) ancestry-specific PRS + non-PRS risk factors, and (3) trans-ancestry PRS + non-PRS risk factors. We measured effect size estimates per PRS unit, P-value significance (<0.05) and discriminative accuracy using area under the receiver operator curve (AUC).[19]

**5. Ancestry-specific principal components in Penn Medicine BioBank**

Penn Medicine BioBank (PMBB) individuals were grouped by their genetically inferred ancestry and principal components were computed after filtering for minor allele frequency $\geq$ 0.05, Hardy-Weinberg equilibrium exact test p-value $\geq 10^{-6}$, and LD-pruning using a window size of 50 kb and an $r^2$ threshold of 0.5.

**6. Phenotypes extracted for phenome-wide and laboratory parameter association study**

We included PennMedicine BioBank (PMBB) participants who underwent genotyping and have accessible electronic health record data. For phenotypes, we extracted International Classification of Diseases Ninth Revision (ICD-9) and Tenth Revision (ICD-10) available up to April 2021 and laboratory measurements up to October 2022 from the electronic health record among PMBB participants. ICD-9 and ICD-10 codes were mapped to phecodes (See Methods). Cases for each phecode were defined as individuals with the phecode on two or more dates and controls were defined as individuals without the corresponding phecode (574-576.99). Following laboratory measurements were used for the analysis: aspartate aminotransferase (AST), maximum (U/L), alanine aminotransferase (ALT), maximum (U/L), alkaline phosphatase, maximum (U/L), total bilirubin, maximum (mg/dL), direct bilirubin, maximum (mg/dL), indirect bilirubin, maximum (mg/dL), gamma-glutamyl transferase (GGT), maximum (U/L), total cholesterol, maximum (mg/dL), triglycerides, maximum (mg/dL), low-density lipoprotein (LDL), maximum (mg/dL), high-density lipoprotein (HDL), minimum (mg/dL), hemoglobin A1C, maximum (%), glucose-fasting, maximum (mg/dL), erythrocyte sedimentation rate (ESR), maximum (mm/hr), C-reactive protein (CRP), maximum (mg/L), thyroid-stimulating hormone (TSH), median (mIU/L), estimated glomerular filtration rate (eGFR), minimum (mL/min/1.73m$^2$), urobilinogen, maximum (mg/dL). All values except for urobilinogen (urinary source) were measured in serum.

**7. Fine Mapping**

We used the causal robust mapping method in meta-analysis (CARMA) which is a Bayesian model applied to fine mapping to ascertain potential causal variants.[20] An R package (https://github.com/ZikunY/CARMA) was applied to determine credible sets using the trans-ancestry summary statistics, using the 1000 Genomes Reference. For each GWAS locus, the posterior probabilities of each variant being a causal variant were computed. At each locus, 95% credible sets comprised of the smallest number of variants with cumulative posterior probability >95% of containing a true causal variant were constructed. From each credible set at GWAS locus, the SNP with highest posterior inclusion probability (PIP) was identified (Supplemental Table 12).

**8. Tissue-specific heritability enrichment analysis**

We ran stratified LD score regression (LDSR)[21] using LDSC (http://www.github.com/bulik/ldsc) v.1.0.1 with the --h2 flag to estimate the enrichment of SNP-based gallstone heritability in various tissue-specific annotations. The --h2 flag instructs LDSC to calculate "partitioned heritability," thereby computing the amount of gallstone heritability located inside and outside of a given annotation (e.g., open chromatin in pancreas). The annotations were based on specifically expressed genes and chromatin states which were created previously using publicly available gene expression and chromatin data.[22] P-values were calculated using the --print-coefficients flag and calculating the one-sided probability of the returned coefficient z-score.

The baseline model LD scores, plink filesets, allele frequencies and variants weights files for the European 1000 genomes project phase 3 were downloaded from https://console.cloud.google.com/storage/browser/broad-alkesgroup-public-requester-pays/LDSCORE. The set of HapMap3 SNPs was used as the variant reference as suggested by the LDSR authors. The tissue-specific annotations were also downloaded from the above link (LDSC_SEG_ldscores/Multi_tissue_gene_expr_1000Gv3_ldscores.tgz and LDSC_SEG_ldscores/Multi_tissue_chromatin_1000Gv3_ldscores.tgz). As described previously,[22] we ran LDSR for each annotation conditioned on both the baseline model and a control model which was created from all genes, as opposed to tissue-specific genes. The control models were included with the tissue-specific annotations that we downloaded. The P-values for coefficients across all annotations were corrected for multiple testing using a false discovery rate (FDR) Q-value method[Q],[23,24] and annotations were considered significantly enriched if they passed an FDR of 5%.

**9. Annotation to chromatin data**

SNPs were annotated based on chromatin data from the HepG2 cell line, hepatocyte-like cells (HLCs) derived from induced pluripotent stem cell (iPSC), human liver samples, colon derived organoids (colonoids), and single-cell data from the pancreas. Chromatin regions were defined with three experiment types: (1) open chromatin regions from ATAC-seq (HepG2, HLC, human liver, colonoids, pancreas), (2) promoter-interacting regions (PIR) from Capture-C (HepG2, HLCs, colonoids) and Hi-C (pancreas), and (3) enhancer-associated histone modifications from ChIP-seq (HepG2). Using R v4.4.0, coordinates were lifted over from hg38 to hg19 using rtracklayer v1.64.0 and intersected with annotations using GenomicRanges v1.56.0. The ATAC-seq/Capture-C data for HLCs was previously reported in Ramdas et al.,[25] the ATAC-seq/Capture-C data for colonoids was previously described in Lasconi et al.,[26] and the snATAC-seq/Hi-C data for pancreas was previously described in Su et al.[27]

<u>ATAC-seq</u>

HepG2: Live HepG2 cells were harvested via trypsinization, followed by a series of DPBS wash steps. 50,000 cells from each sample were pelleted at 550 ×g for 5 min at 4°C. The cell pellet was then resuspended in 50 µl cold lysis buffer (10 mM Tris-HCl, pH 7.4, 10 mM NaCl, 3 mM MgCl2, 0.1% IGEPAL CA-630) and centifuged immediately at 550 ×g for 10 min at 4°C. The nuclei were resuspended in the transposition reaction mix (2x TD Buffer (Illumina Cat #FC-121-1030, Nextera), 2.5ul Tn5 Transposase (Illumina Cat #FC-121-1030, Nextera) and Nuclease Free $H_2O$) on ice and then incubated for 45 min at 37°. The transposed DNA was then purified using the MinElute Kit (Qiagen), eluted with 10.5 µl elution buffer (EB). The transposed DNA was PCR amplified using Nextera primers for 12 cycles to generate each library. The PCR reaction was subsequently cleaned up using AMPureXP beads (Agencourt) and libraries were paired-end sequenced on an Illumina Novaseq 6000 (51 bp read length). ATAC-seq for colonoid, HLC, and HepG2 cells was processed using the ENCODE ATAC-seq pipeline. IDR optimal peaks were selected as the consensus peakset per celltype. The human liver ATAC-seq samples will be available pending the publication of a manuscript by M.F.D. Open chromatin peaks were called on all 189 human liver samples using Genrich (https://github.com/jsh58/Genrich) with parameters -j, -m 10, -p 1e-40, -a 3000, and -g 50.

Capture-C

Capture-C data for all cell types was prepared and analyzed as in Chesi et al.[28] Briefly Capture C sequencing data was processed using the HiCUP pipeline (v 0.7.4), which aligned reads to hg19 using bowtie2 (v2.2.6) and remove duplicate and invalid reads.[29] Following this, chromatin contacts were called using Chicago (v1..1.8)[30] with the bait and restriction enzyme maps for DpnII and prior reported bait file.[28] Our capture baits were designed to target both ends of the *DpnII* restriction fragments containing promoters for coding mRNA, non-coding RNA, antisense RNA, snRNA, miRNA, snoRNA, and lincRNA transcripts (UCSC lincRNA transcripts and sno/miRNA under hg19 assembly) totaling 36,691 RNA baited fragments through the genome. Interactions were called using either individual restriction fragment (1-frag), or with four consecutive fragments concatenated in silico (4-frag). Chicago was run with the default parameters except with the binSize set to 2500 for 1frag and removeAdjacent set to False for 4frag.

“Promoter-interacting regions” were defined as CHiCAGO peaks with a score (distance adjusted log transformed P value) ≥ 5, which appeared using either 1-fragment or 4-fragment resolution.

Histone ChIP-seq

HepG2 Cells were fixed in the same manner as described in the Promoter Capture methods. Fixed cells were then subjected to sonication following the NEXSON procedure.[31] Briefly, nuclei were isolated by sonication using a Q Sonica Sonicator with settings of 15 seconds On, 30 seconds Off for 8 cycles at 20% power (2 minutes total sonication time). Nuclei were centrifuged at 1250 RCF and resuspended in CHIP Buffer. DNA was sheared using sonicator settings of 30 seconds On, 30 seconds Off at 80% power for 4 cycles of 5 minutes each (20min. total sonication time). Samples were centrifuged at 18,000 RCF and supernatant was collected and tested on an agarose gel to check for size, phenol extracted, precipitated, and resuspended. ChIP was carried out with 300ng sheared DNA and the Chip-IT High Sensitivity Kit (Active Motif, 53040) according to kit instructions with the following antibodies from Active Motif: H3K27ac (39685), H3K4me1 (39297), and H3K4me3 (61379). Libraries were generated from the resulting material using the ThruPLEX Tag-seq Library Kit (Takara, 030520), PCR amplified for 10-16 cycles then purified and size selected using AMPure XP Beads. Completed libraries were checked for quality and quantity on a BioAnalyzer 2100 and Qubit respectively. Libraries passing QC were pooled at 1nM and sequenced on a Novaseq 6000 using an S1 flow cell (51bp paired end read length, dual indexed).

**10. Cross-trait colocalization**

Colocalizations were performed between the multi-ancestry gallstone GWAS and 44 related trait GWAS.[6,32–45] Before colocalization, all SNPs were assigned rsIDs based on dbSNP build 155, which were used for matching across traits. For each lead variant in gallstone, a 500kb window centered on the window was colocalized if a variant in the window had a -value < $1x10^{-5}$. Of 1511 trait pair regions tested, 350 had a positive colocalization result, defined as a PP4 (posterior probability for sharing trait pair signals) > 0.8. Colocalization was tested using COLOC 5.1.0[46] using beta, varbeta ($SE^2$), sample sizes, gallstone MAF, and if appropriate, case control ratios. The top SNP in each region was identified from the colocalization output and used to determine effect direction concordance.

In order to leverage the higher power available for LDL, TBil and DBil GWAS, we also tested for colocalization all gallstone loci with a lead p-value x, where $5x10^{-5} > x > 5 \times 10^{-8}$. This tested a further 426 loci and yielded another 16 colocalizations.

**11. Expression quantitative trait locus (eQTL) colocalization**

We selected the GTEx tissues colon-sigmoid, colon-transverse, liver, pancreas, small intestine-terminal ileum, spleen, and whole blood to colocalize with multi-ancestry gallstone GWAS. The summary statistics from GTEx v8 for each tissue were used for the analysis.[47] We used the Colocquial pipeline[48] to test for colocalization, testing 343 gene/tissue pairs and identifying 97 positive colocalizations using the threshold PP4 > 0.8. The top SNP in each region was identified from the colocalization output and used to determine effect direction concordance. We additionally tested regions that colocalized between gallstone and LDL, TBil or DBil for colocalization between the non-gallstone trait and the above GTEx tissues using the same methods.

**12. Gene set analysis**

We performed gene set analysis using the Functional Mapping and Annotation (FUMA) platform.[49] We used the GENE2FUNC function to associate the nominated genes of 91 lead SNPs from trans-ancestry and ancestry-specific analyses with biological pathways from KEGG, Reactome, Wikipathways, and Gene Oncology.[49] Specifically, overlaps between the nominated genes and the gene sets related to biological pathways were tested using hypergeometric tests.[49] We excluded the MHC region from the analysis given its high pleiotropy.[50] We reported gene sets of biological pathways with adjusted P value <0.05 using Benjamini-Hochberg false discovery rates.[49]

#### 13. Supplemental figures

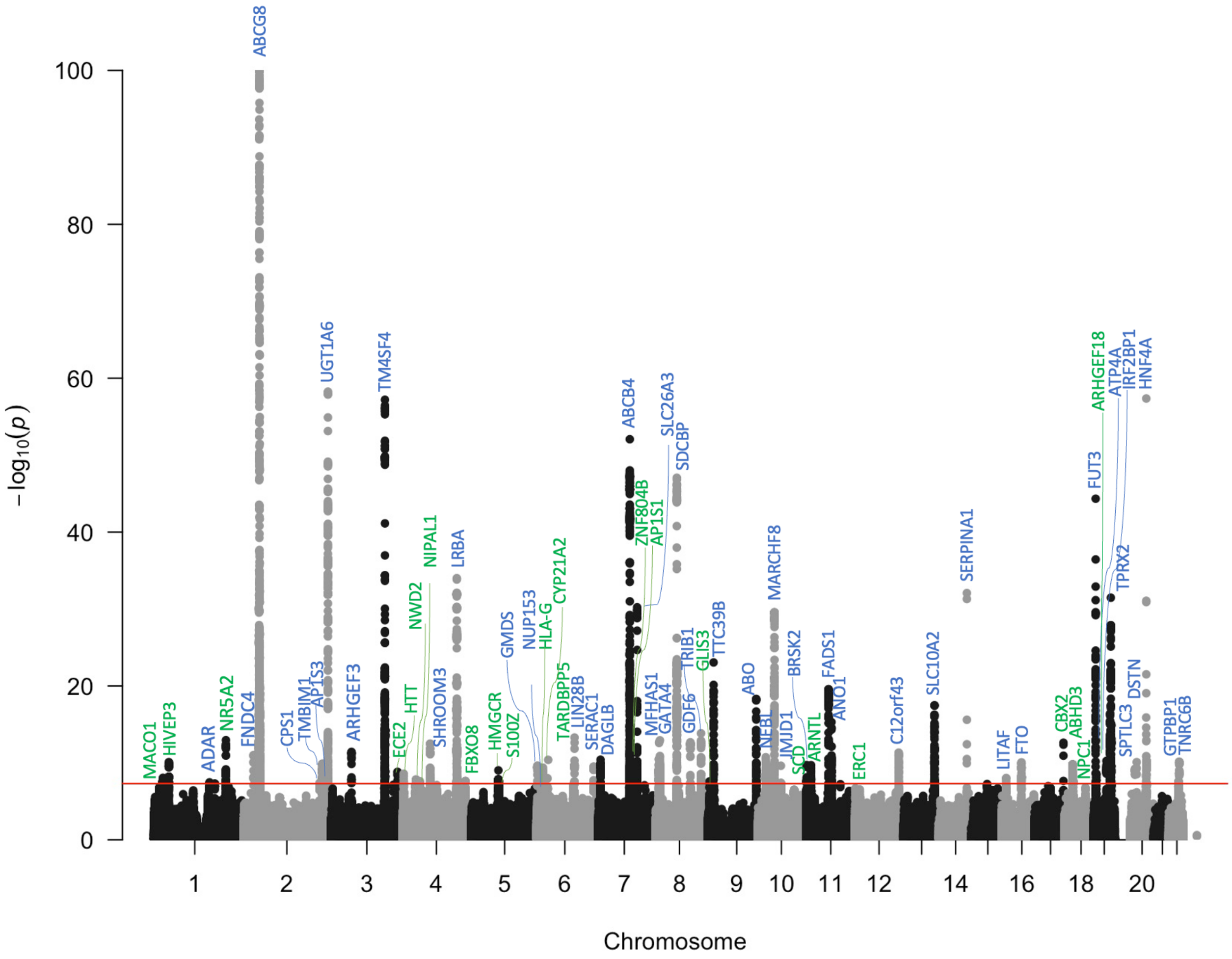


**Figure S1. Manhattan plot of genome-wide association meta-analysis in European ancestry** (72,827 gallstone cases and 1,158,486 controls). 68 loci met the genome-wide significance ($p = 5 \times 10^{-8}$), represented by red line. Novel loci are annotated in green. Known loci are annotated in blue. Y-axis is truncated at $p = 10^{-100}$

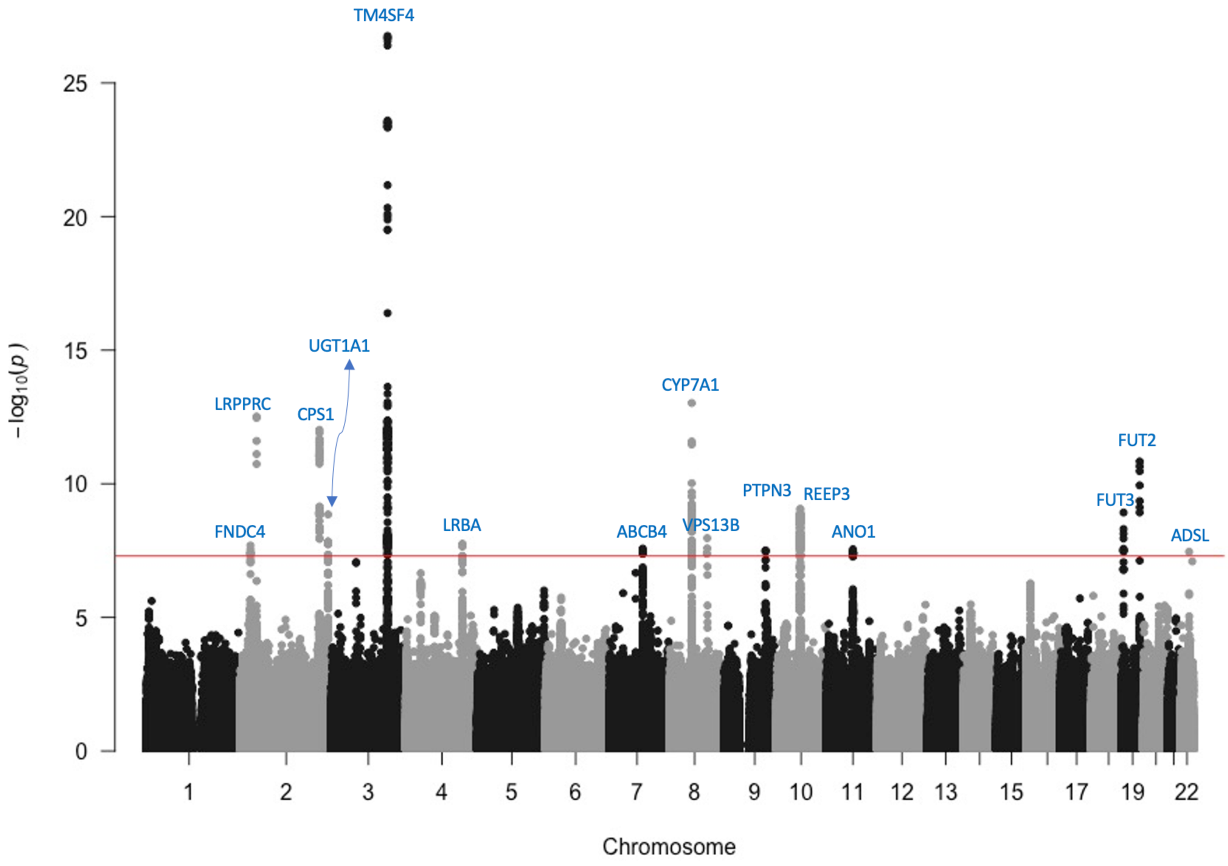


**Figure S2. Manhattan plot of genome-wide association study in East Asian ancestry** (9,305 gallstone cases and 168,253 controls). 15 loci met the genome-wide significance (p = 5 x $10^{-8}$), represented by red line. Novel loci are annotated in blue.

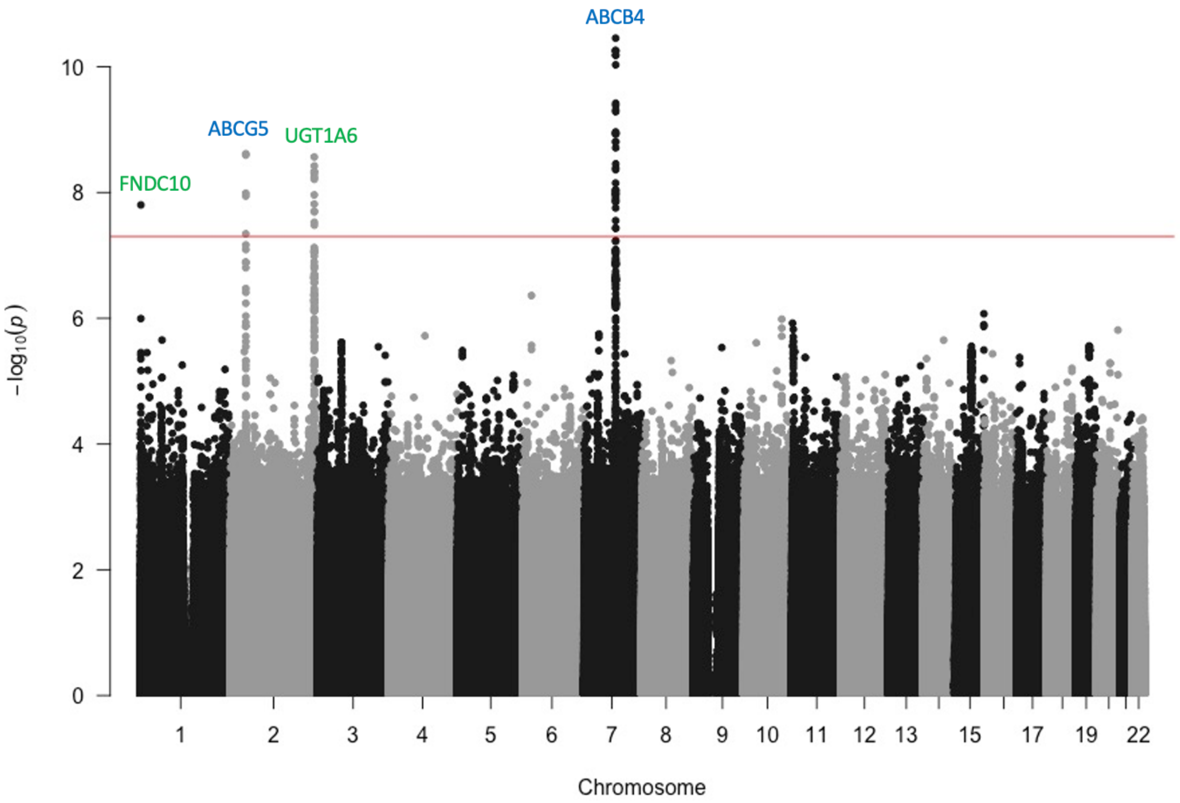


**Figure S3. Manhattan plot of genome-wide association study in African ancestry** (3,636 gallstone cases and 115,711 controls). 4 loci met the genome-wide significance (p = 5 x $10^{-8}$), represented by red line. Novel loci are annotated in blue.

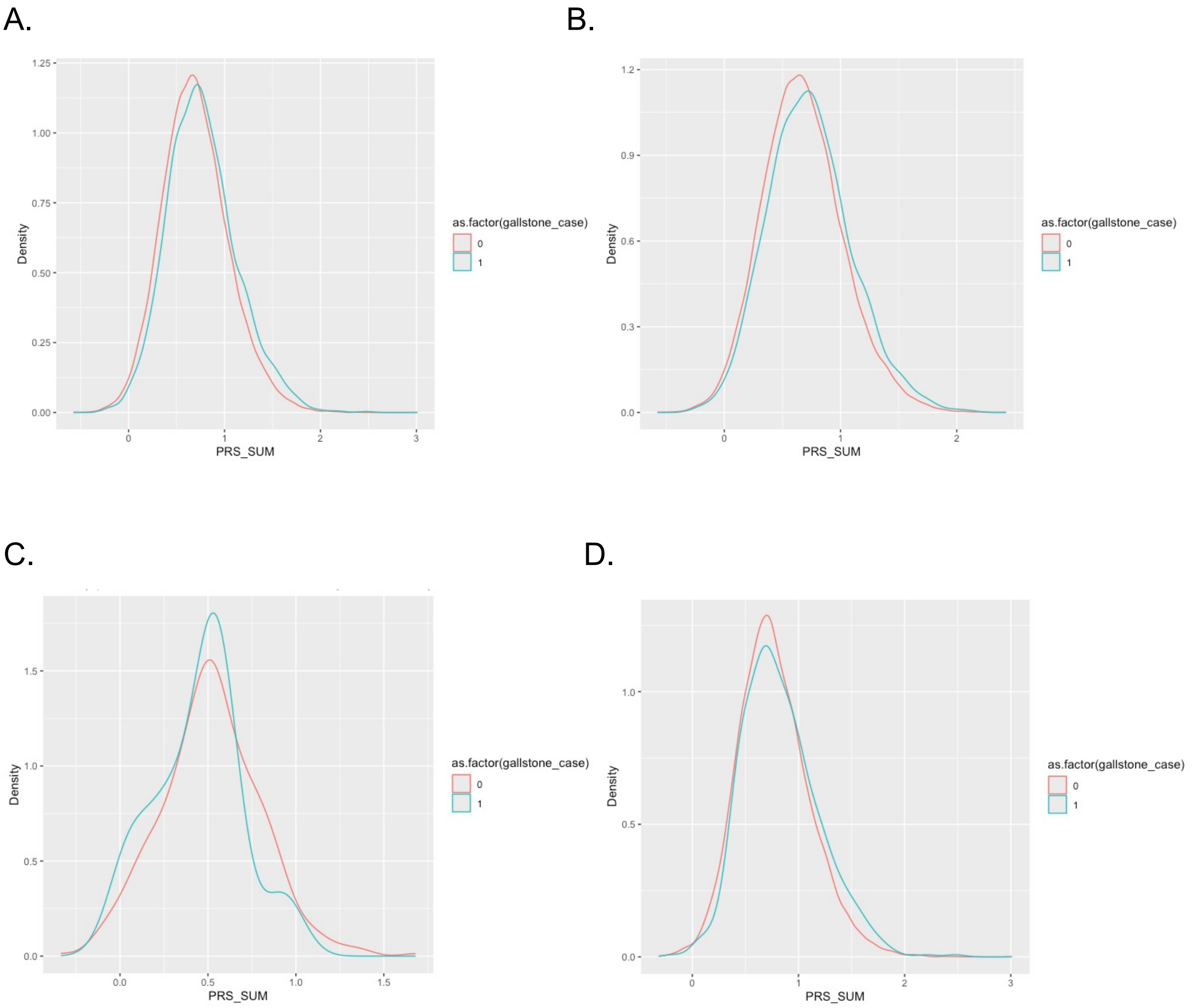


**Figure S4. Density plots of gallstone cases in Penn Medicine BioBank by trans-ancestry polygenic risk score constructed using weights based on the risk allele effects for lead SNPs. A.** Gallstone cases in trans-ancestry cohort. **B.** Gallstone cases among those of European ancestry. **C.** Gallstone cases among those East Asian ancestry. **D.** Gallstone cases among those of African ancestry.

A. B.

C.

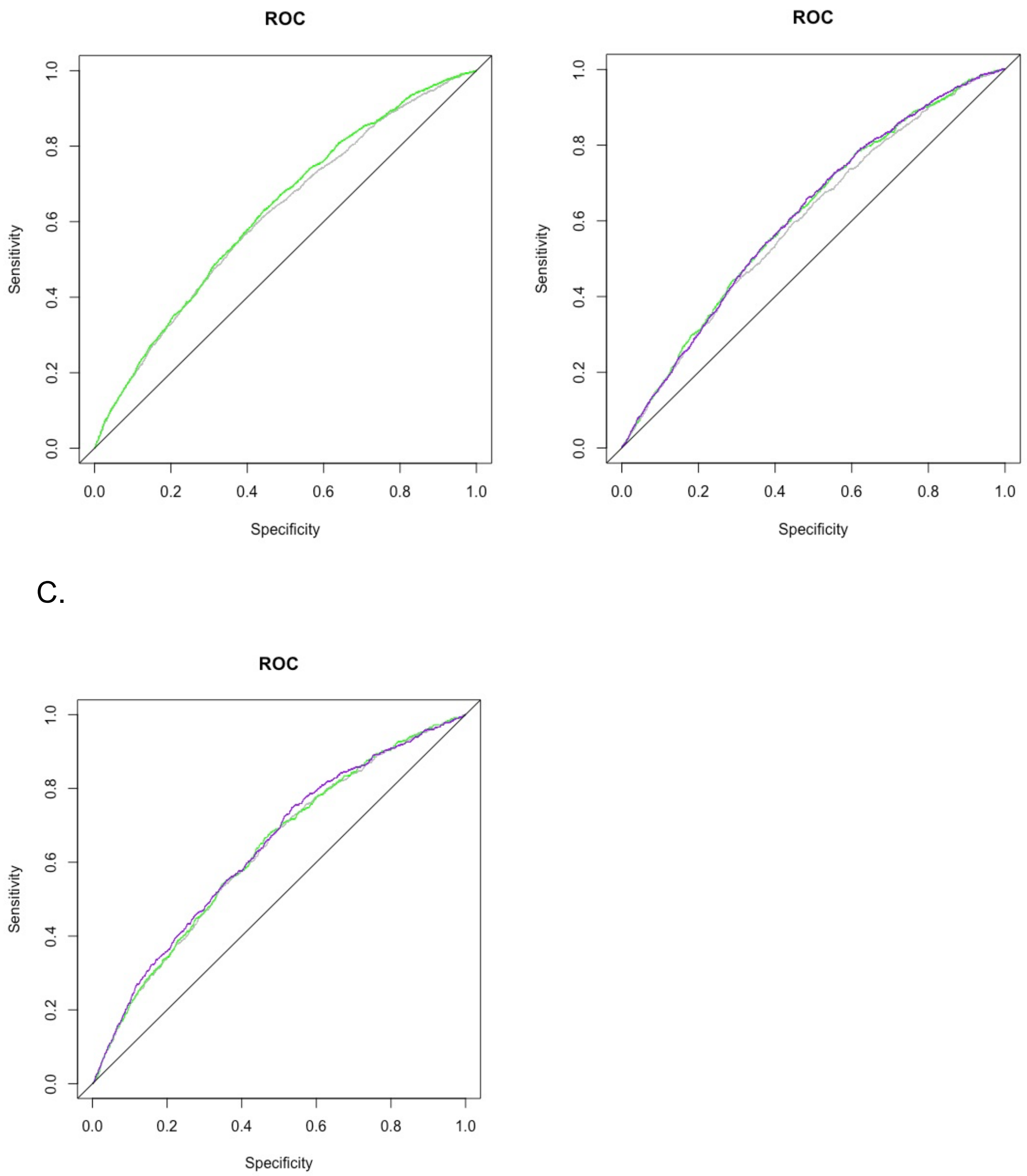


**Figure S5. Receiver operating characteristic (ROC) plot of polygenic risk score (PRS) in Penn Medicine BioBank**

**A.** Trans-ancestry PRS + non-PRS risk factors (Green; AUC = 62.7%), non-PRS risk factors (Grey; AUC = 61.6%) in trans-ancestry group. **B.** Trans-ancestry PRS + non-PRS risk factors (Purple; AUC = 61.4%), European ancestry PRS + non-PRS risk factors (Green; AUC = 61.3%), non-PRS risk factors (Grey; AUC = 60%) in European-ancestry group. **C.** Trans-ancestry PRS + non-PRS risk factors (Purple; AUC = 63.8%), African ancestry PRS + non-PRS risk factors (Green; AUC = 63.1%), non-PRS risk factors (Grey; AUC = 62.8%) in African ancestry group.

*Reference line (Black; AUC = 50%)

*non-PRS risk factors: age, genetically inferred sex, body mass index [median], and first 10 principal components of genetic ancestry

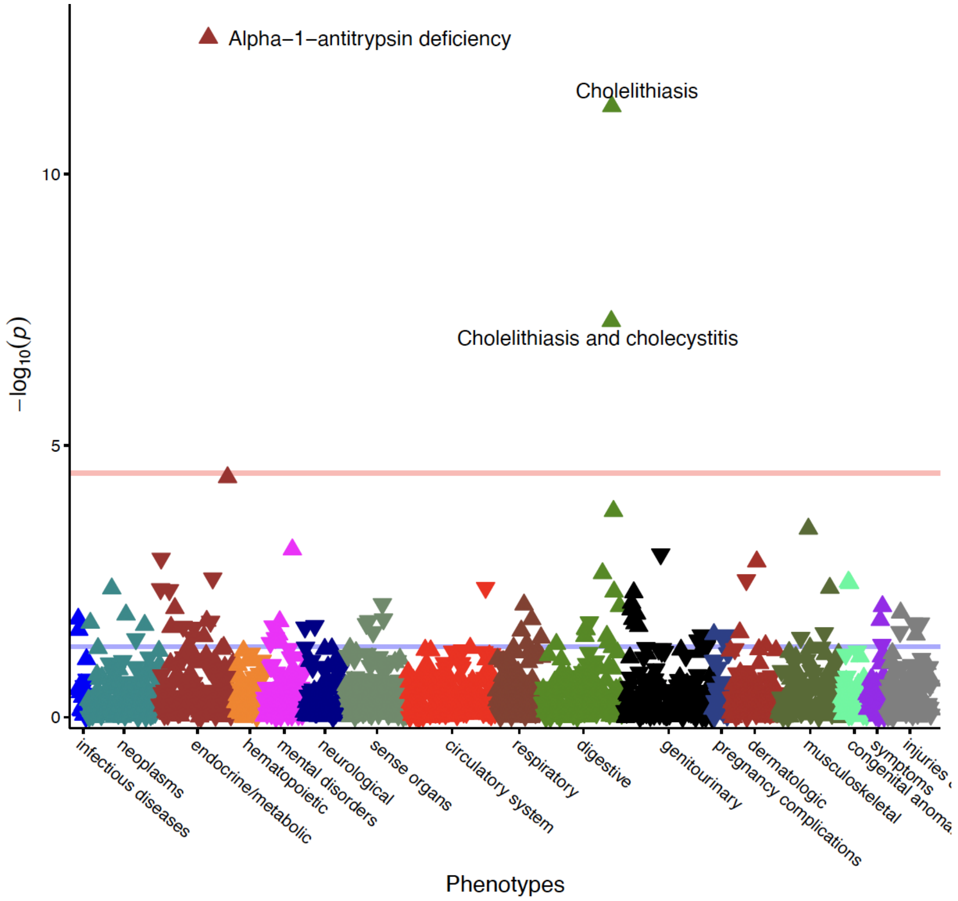


**Figure S6. Phenome-wide association study of trans-ancestry polygenic risk score in trans-ancestry cohort in Penn Medicine BioBank.** Purple line indicates P-value of 0.05. Orange value indicates Bonferroni adjusted P-value of 2.86 x$10^{-5}$.

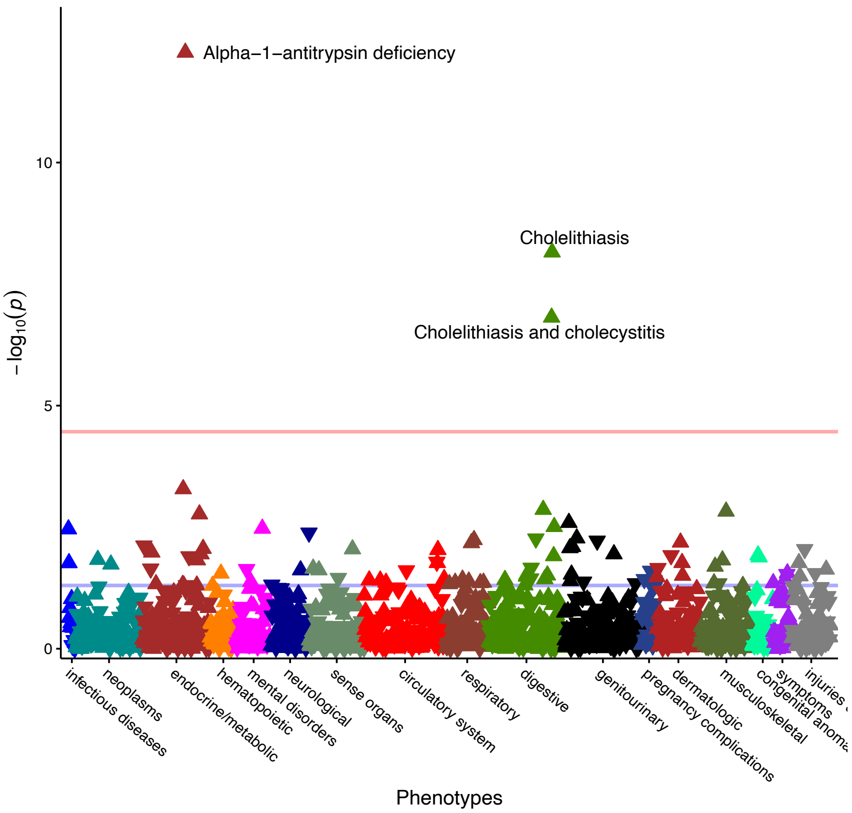


**Figure S7. Phenome-wide association study of trans-ancestry polygenic risk score in European ancestry cohort in Penn Medicine BioBank.** Purple line indicates P-value of 0.05. Orange value indicates Bonferroni adjusted P-value of 2.862 $x10^{-5}$.

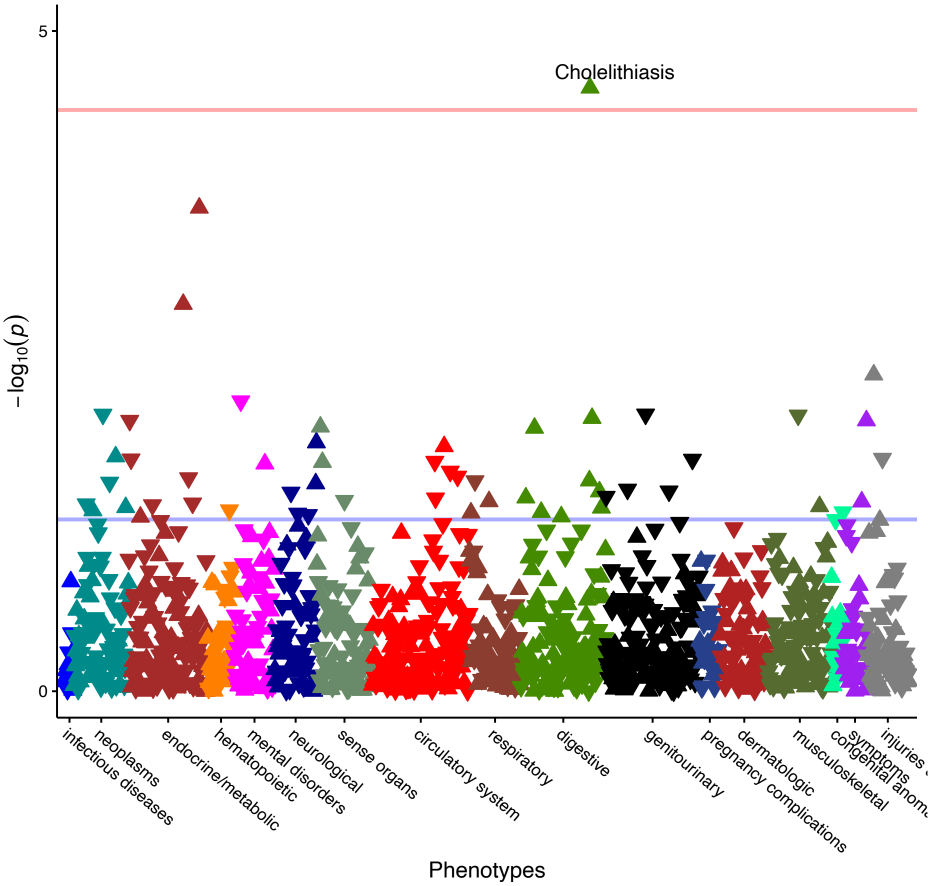


**Figure S8. Phenome-wide association study of trans-ancestry polygenic risk score in African ancestry cohort in Penn Medicine BioBank.** Purple line indicates P-value of 0.05. Orange value indicates Bonferroni adjusted P-value of 2.862 $\times 10^{-5}$.

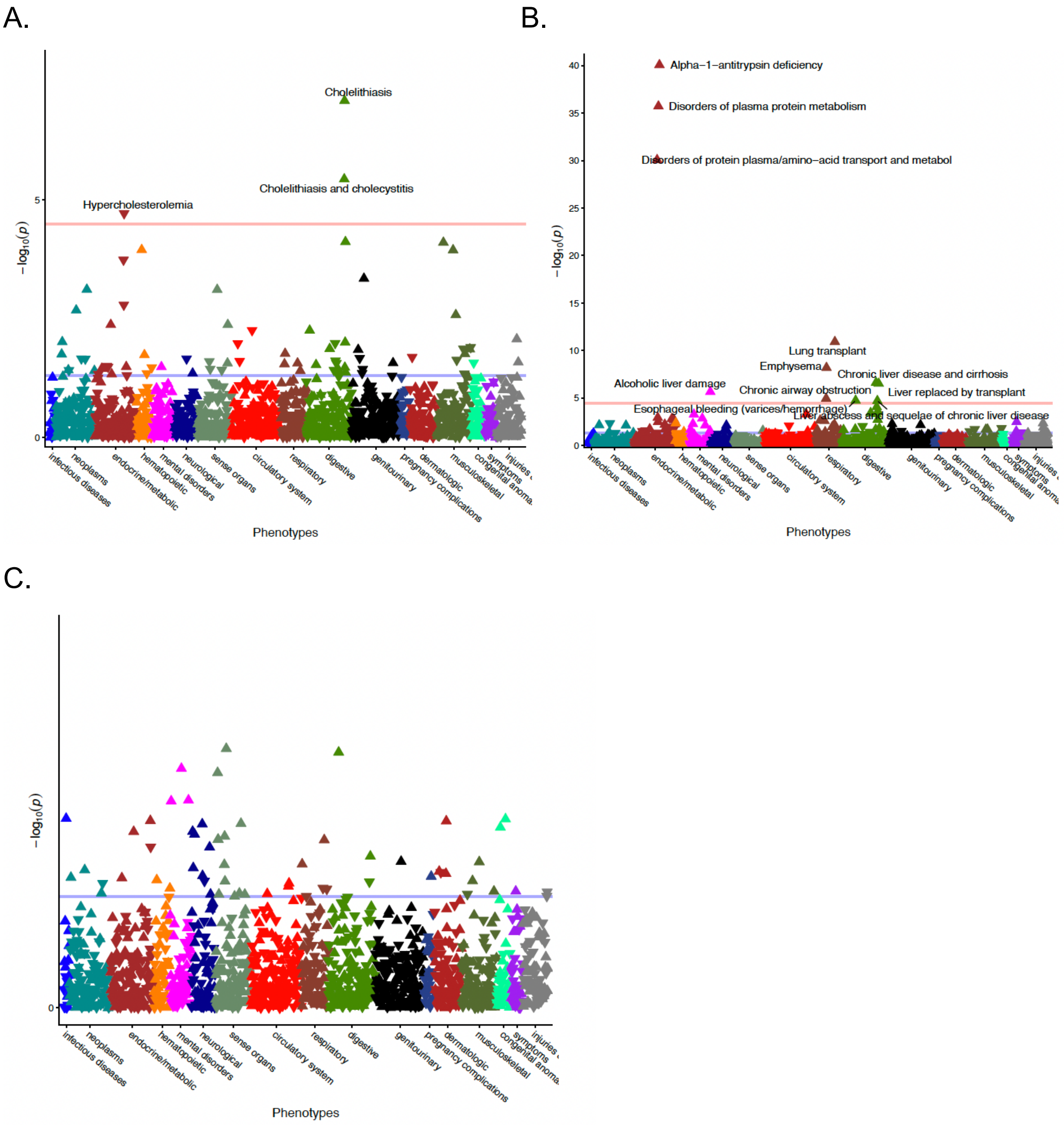


**Figure S9. Phenome-wide association study of missense coding variants in trans-ancestry cohort in Penn Medicine BioBank.** Purple line indicates P-value of 0.05. Orange value indicates Bonferroni adjusted P-value of 2.862 x10$^{-5}$. **A.** rs11887534, ABCG8 (gene nominated through Open Targets Genetics). **B.** rs28929474, SERPINA1 (gene nominated through Open Targets Genetics). **C.** rs1800961, HNF4A (gene nominated through Open Targets Genetics)

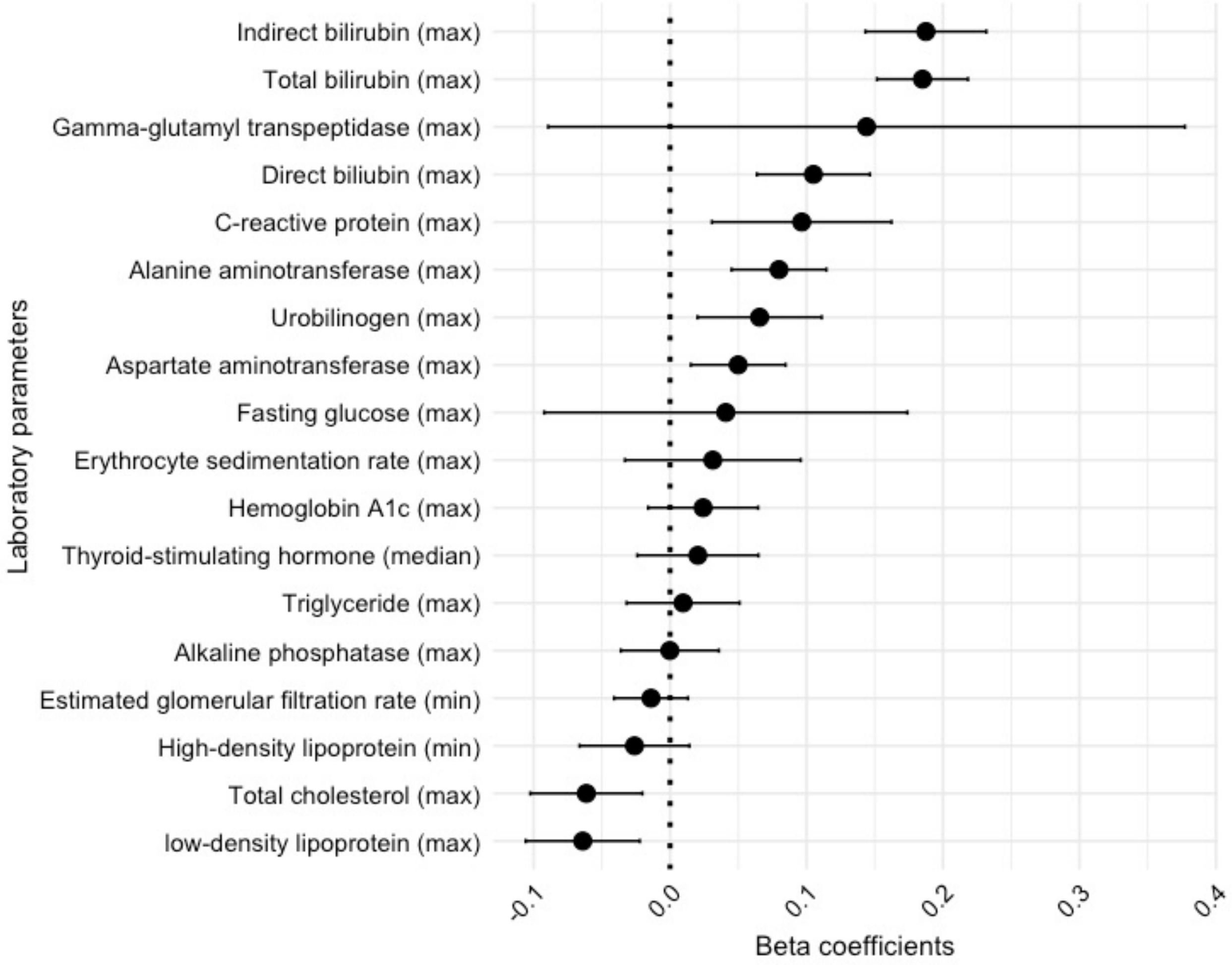


**Figure S10. Laboratory parameter association study of the trans-ancestry polygenic risk score in the Penn Medicine BioBank European ancestry cohort.** Linear regression was used to test the associations between laboratory measurements extracted from the PMBB-linked electronic health record and the trans-ancestry polygenic risk score constructed in PMBB. Bonferroni-significant threshold was set at $P < 2.778 \times 10^{-3}$.

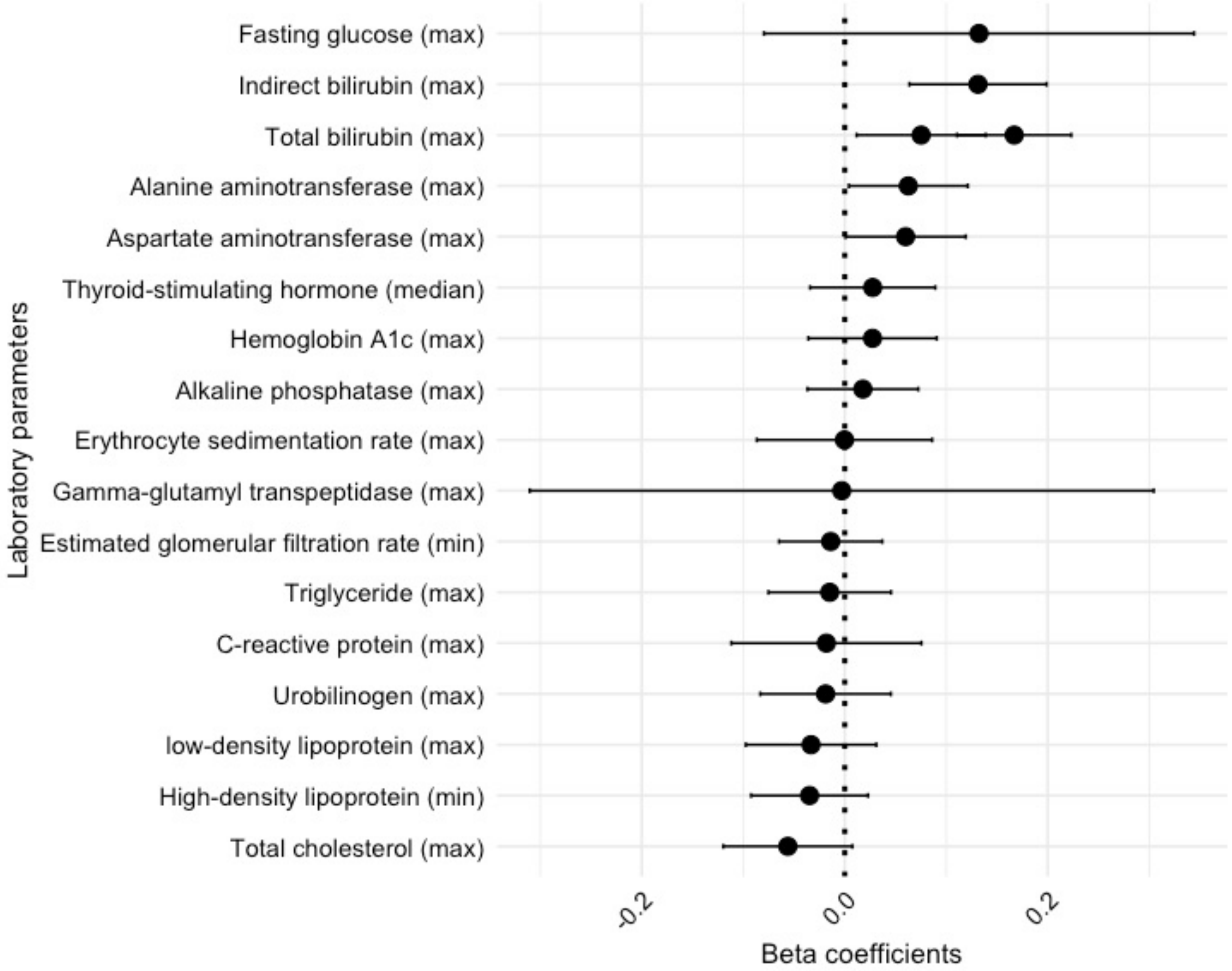


**Figure S11. Laboratory parameter association study of the trans-ancestry polygenic risk score in the Penn Medicine BioBank African ancestry cohort.** Linear regression was used to test the associations between laboratory measurements extracted from the PMBB-linked electronic health record and the trans-ancestry polygenic risk score constructed in PMBB. Bonferroni-significant threshold was set at $P < 2.778 \times 10^{-3}$.

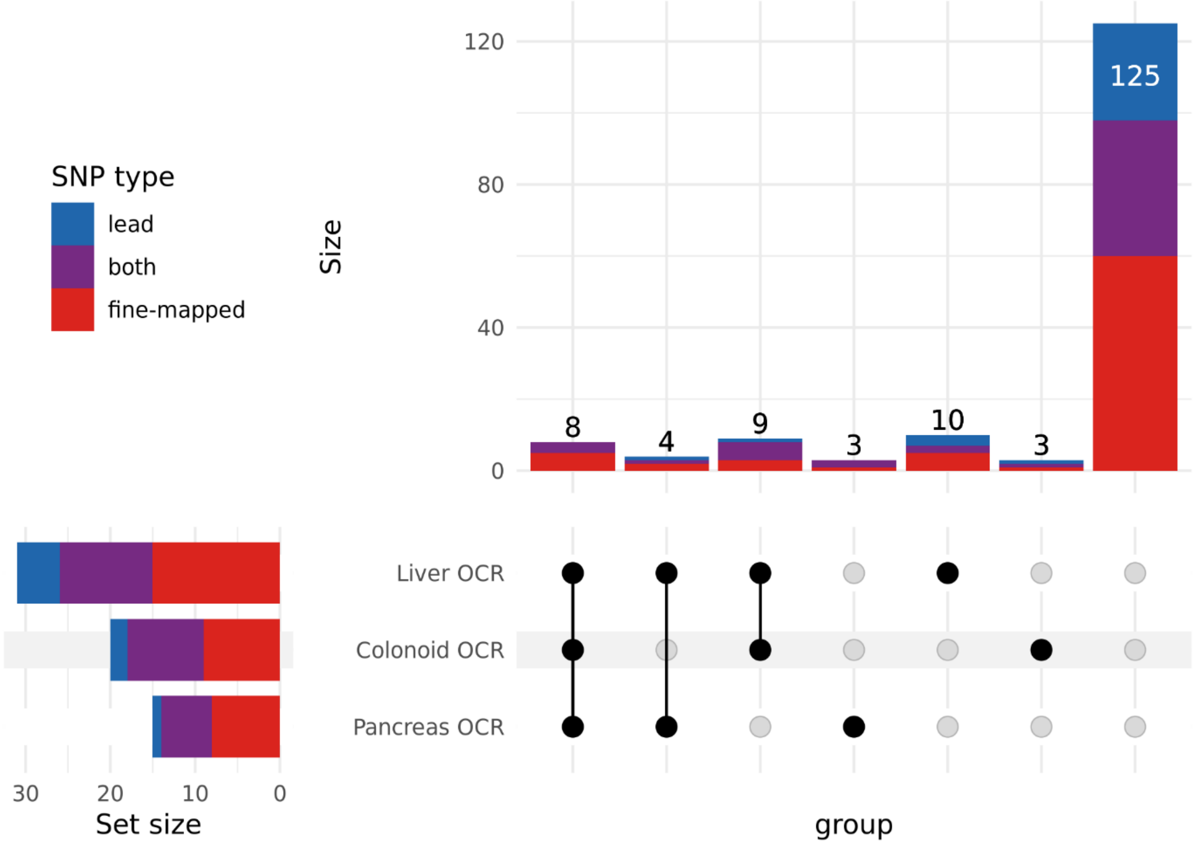


**Figure S12. UpSet plot of the overlap between loci identified in trans-ancestry meta-analysis and open chromatin regions in liver, colonoid, and pancreas tissue.** Variants in Liver OCR were open in any of the liver ATAC cell types (HepG2, HLC, or human liver). Variants in Pancreas OCR were open in any of the pancreatic ATAC cell types (alpha, acinar, beta, delta, ductal, endothelial, epsilon, or pancreatic polypeptide).

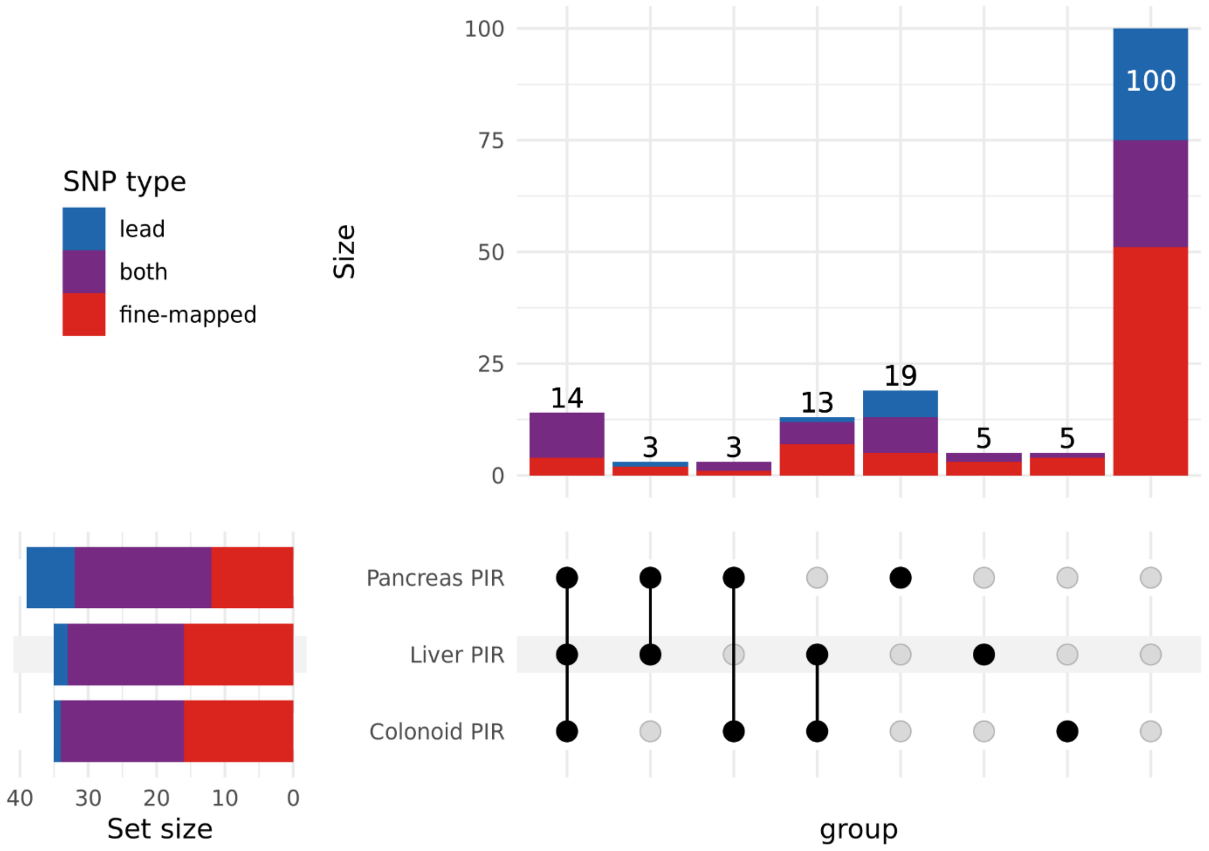


**Figure S13. UpSet plot of the overlap between loci identified in trans-ancestry meta-analysis and promoter interacting regions in liver, colonoid, and pancreas tissue.** Variants in Liver PIR were open in any of the liver Capture-C cell types (HepG2 or HLC). Variants in Pancreas OCR were open in any of the pancreatic Hi-C cell types (alpha, acinar, or beta).

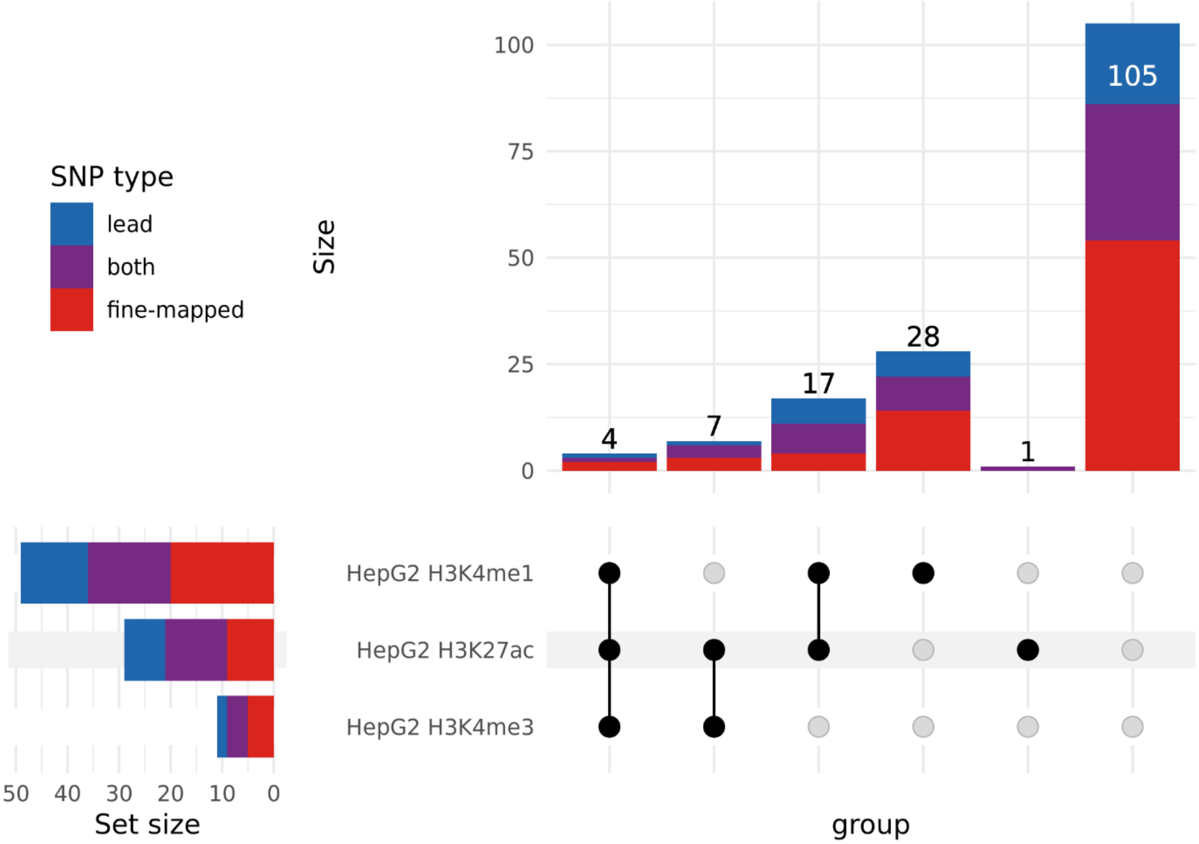


**Figure S14. UpSet plot of the overlap between loci identified in trans-ancestry meta-analysis and histone marks in HepG2.**

**14. Penn Medicine BioBank Banner Author List and Contribution Statements**

Penn Medicine BioBank Banner Author List includes following:

PMBB Leadership Team

- Daniel J. Rader, MD, Marylyn D. Ritchie, PhD.
- Contribution: All authors contributed to securing funding, study design and oversight. All authors reviewed the final version of the manuscript.

Patient Recruitment and Regulatory Oversight

- JoEllen Weaver, Nawar Naseer, PhD, MPH, Giorgio Sirugo, MD, PhD, Afiya Poindexter, Yi-An Ko, PhD, Kyle P. Nerz
- Contributions: JW manages patient recruitment and regulatory oversight of study. NN manages participant engagement, assists with regulatory oversight, and researcher access. GS assists with researcher access. AP, YK, KPN perform recruitment and enrollment of study participants.

Lab Operations

- JoEllen Weaver, Meghan Livingstone, Fred Vadivieso, Stephanie DerOhannessian, Teo Tran, Julia Stephanowski, Salma Santos, Ned Haubein, P.h.D., Joseph Dunn
- Contribution: JW, ML, FV, SD conduct oversight of lab operations. ML, FV, AK, SD, TT, JS, SS perform sample processing. NH, JD are responsible for sample tracking and the laboratory information management system.

Clinical Informatics

- Anurag Verma, PhD, Colleen Morse Kripke, MS, DPT, MSA, Marjorie Risman, MS, Renae Judy, BS, Colin Wollack, MS
- Contribution: All authors contributed to the development and validation of clinical phenotypes used to identify study subjects and (when applicable) controls.

Genome Informatics

- Anurag Verma PhD, Shefali S. Verma, PhD, Scott Damrauer, MD, Yuki Bradford, MS, Scott Dudek, MS, Theodore Drivas, MD, PhD,
- Contribution: AV, SSV, and SD are responsible for the analysis, design, and infrastructure needed to quality control genotype and exome data. YB performs

the analysis. TD and AV provides variant and gene annotations and their functional interpretation of variants.
